# Making inferences with incomplete epidemiological data: a proof-of-concept estimating measles vaccine coverage across Canada

**DOI:** 10.64898/2026.07.16.26358151

**Authors:** Liza Hadley, Rachael M. Milwid, Valerie Hongoh, Rania Wasfi, Stephen M. Kissler, Irena Papst

## Abstract

**Background & aims of study:** Current measles outbreaks around the world have highlighted the evolving immunity landscape of some vaccine-preventable diseases (VPDs). In Canada, there have been approximately 1,100 reported measles cases between January and June 2026 alone. This far exceeds the previous average annual case counts of <200. This significant outbreak has underscored the need to better understand the present state of population immunity to measles in Canada, an essential input parameter for outbreak response models. Most population immunity in this setting is derived from routine childhood vaccination, but vaccine coverage data is available heterogeneously across the country and is typically collected cross-sectionally to monitor population adherence to immunization schedules rather than to inform population-level susceptibility. This study developed statistical methods to adapt available measles vaccine coverage estimates into more complete estimates of present-day immunity in Canada by age and province/territory (PT).

**Methods & results:** First, a standardized dataset for vaccine coverage by PT was collated from existing publicly-available datasets and reports. A significant number of vaccine coverage estimates were missing by PT, dose, and year. We used Gaussian Process models to impute current coverage with at least one dose of a measles vaccine, to create a complete ‘modelled dataset’ by PT and age. Our modelling framework was tested against data for England, which was much more complete, to validate our approach.

**Implications:** We developed methodology for estimating vaccine coverage in the absence of detailed population immunity data, to support ongoing measles outbreak modelling response work. While this project focused on measles, we built the associated code/tools such that the methodology can be applied to other vaccine-preventable diseases in future outbreak settings.

**Highlights:**

- Using statistical modelling, we estimated current measles vaccine coverage in Canada.
- We developed a flexible Gaussian Process model as a proof of concept.
- Regions were related using socioeconomic and vaccine hesitancy indicators.

## 1 Introduction

Measles has been an important cause of child morbidity and mortality over the last century, owing in large part to its highly contagious nature and respiratory route of transmission [1,2]. Despite this, the availability of a safe and effective live-attenuated vaccine has allowed a number of countries to achieve measles elimination status, including Canada which first achieved this status in 1998 [3,4]. Most outbreaks in Canada between 1998 and 2024 were transient and contained, originating from imported cases from measles endemic regions [5]. However, Canada’s measles elimination status was revoked in November 2025 following a sustained outbreak beginning in October 2024 and lasting more than 12 months [6]. Other countries have also recently lost their elimination status, including the UK, Spain, Austria, Armenia, Azerbaijan, and Uzbekistan, as of September 2025 [7]. The outbreak of measles in Canada has resulted in approximately 6,500 cases as of June 2026, with cases reported in 7/13 of the provinces/territories (PTs) [8]. The majority (98%) of cases from 2025 were linked to an exposure within Canada. Most reported cases (89%) were among unvaccinated individuals, with 4% of cases occurring among people with an unknown vaccination status [8]. 45% of cases in 2025 were among youth aged 5-17 years of age, with 20% of cases occurring in those aged 1-4 and 28% occurring in those between the ages of 18-54 [8].

To re-acquire measles elimination status, it is crucial to develop a national view of the immunity landscape that captures age and regional differences, enabling identification of high-risk population pockets for targeted public health action. Here immunity landscape refers to both naturally-acquired and vaccine-induced immunity. In Canada, publicly-funded routine measles vaccination began in the 1970s [4]; adults born before 1970 are generally assumed to have acquired robust natural immunity from infection, as reflected in the case data where fewer than 1% of cases in 2025 occurred in adults over 55 [8,9]. Immunity in younger cohorts, on the other hand, is typically derived through vaccination due to extremely low prevalence of measles cases in Canada the last four decades [10,11].

Vaccination requirements, schedules, and reporting are mandated at the provincial/territorial level, leading to heterogeneity in the amount of vaccine coverage data across the country (Figure 1). Furthermore, vaccination data is not always publicly-available; to date, there is no central, publicly-available repository that collects and/or standardizes vaccination data from each province and territory in Canada. A new initiative, STARVAX (Standardized Reporting on Vaccination), is starting to collect standardized data, but at present only nine of 13 PTs are participating and PT-stratified data has not been made publicly-available [12,13]. Serological data may present a good alternative to vaccine coverage data in quantifying the measles immunity landscape in Canada, though available studies are either based on samples taken over a decade ago and/or feature data only from a few PTs [14–17].

**Figure 1:**
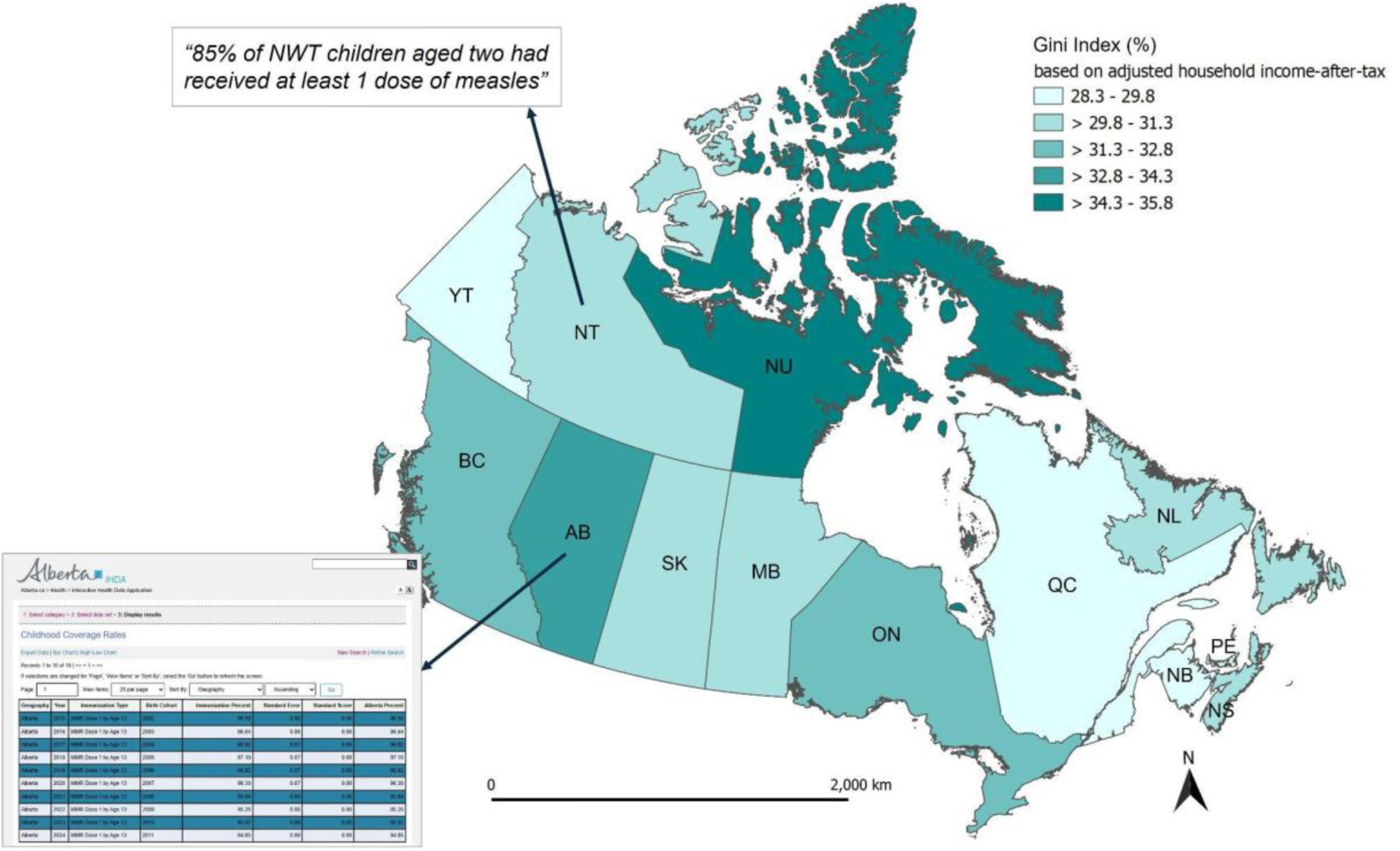
Map of Canada illustrating the heterogeneity in the amount of publicly-available vaccine coverage data between provinces/territories. Some PTs have rich vaccine coverage data dashboards (e.g. Alberta, bottom left [18]) while others have minimal publicly-available reporting (e.g. Northwest Territories, upper left [19]). The map is additionally coloured to show each PT’s Gini index to illustrate Canada’s additional socioeconomic heterogeneity among PTs. The Gini index shown here is calculated based on adjusted household income-after-tax (mean of 2015 and 2020), and measures income inequality with a coefficient that ranges from 0% (complete equality) to 100% (complete inequality) [20]. Abbreviations are as follows: British Columbia (BC), Alberta (AB), Saskatchewan (SK), Manitoba (MB), Ontario (ON), Quebec (QC), Newfoundland and Labrador (NL), New Brunswick (NB), Nova Scotia (NS), Prince Edward Island (PE), Yukon (YT), Nunavut (NU), Northwest Territories (NT/ NWT). Map uses the NAD83 Statistics Canada Lambert Conformal Conic projected coordinate system [21].

The absence of publicly-available vaccine coverage data by age and region and of serosurveys with higher population coverage in Canada, coupled with the ongoing outbreak and recent loss of measles elimination status, has identified a pressing need to find alternative methods for quantifying the population’s susceptibility to measles. Settings outside of Canada have faced the similar problem of needing to estimate measles vaccine coverage from incomplete data [22–26]. For example, Kumar et al. developed a step-wise joint Bernoulli model incorporating between-dose correlation and following specific birth cohorts in Malaysia [25], while Bhatia et al. developed a beta regression model using routine surveillance data to predict subnational coverage in measles-endemic countries in Africa [24]. Utazi et al. and Sbarra et al. both utilised Gaussian Process models for low-and-middle-income settings but took a more computationally-intensive approach [22,23]. More similar to our own setting but with more comprehensive datasets, a research group in North Carolina have been able to make predictions of the number of individuals vaccinated against measles at the county level in the US from count data [26]. Here, we explored a statistical modelling method, Gaussian Process (GP) models, based on vaccine coverage data.

We developed GP models as a proof of concept for estimating current measles vaccination coverage by age and region, where data availability is patchy, as in Canada. Given the sparsity of the Canadian vaccine coverage data, it was not possible to directly validate our estimates. Instead, we constructed a comparable model within the same framework using subsets of more complete data from England, a country with similar socioeconomic status and healthcare access (largely free at point-of-use) to Canada, to help validate our methodology. Using the GP approach, we considered four indicators to relate vaccination coverage among different PTs in Canada motivated by literature on social determinants of health: the Gini coefficient, the proportion of children in low income families, vaccine hesitancy among parents (pre-pandemic), and the proportion of children who did not get vaccinated against COVID-19 (in 2022).

## 2 Materials and methods

### 2.1 Collated vaccine coverage data

We first performed an exhaustive search of the academic and grey literature, focusing on provincial/territorial public health pages, for publicly-available Canadian measles vaccine uptake data. We focused on estimates stemming from vaccine administration data, as opposed to serosurveys. Where available, we collected both PT- and sub-PT-level data. In the case that no data were found, a Google search was performed to identify any additional relevant estimates of vaccine uptake from sources including news articles and press releases. Search terms included “measles vaccine coverage + [PT]” and “MMR vaccine coverage + [PT]”. Published national, population-level survey results for children from the Childhood National Immunization Coverage Survey (cNICS) [27] and for 18+ year olds from the Adult National Immunization Coverage Survey (aNICS; 2023) [28] were used to supplement PT-reported data. No publicly-available vaccine coverage data released by the PT could be found for Saskatchewan, New Brunswick, Nova Scotia, Yukon, and Nunavut; here, we use estimates from the national surveys only (cNICS, aNICS).

The raw collated data is summarized in Table 1. All data was collated and standardized to reconcile labelling differences for age cohorts (age at vaccination vs. birth year) and the number of doses received (one dose, two doses, “1+” dose, “2+” doses). The full standardized dataset is available on GitHub (https://github.com/phac-modelling-hub/immunity-landscape).

**Table 1:**
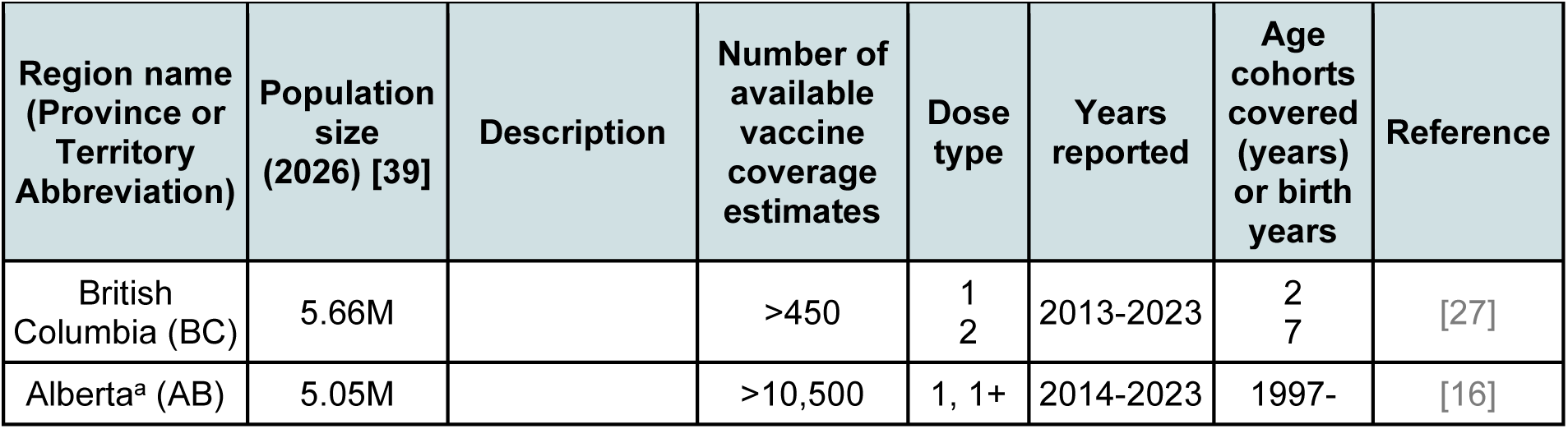
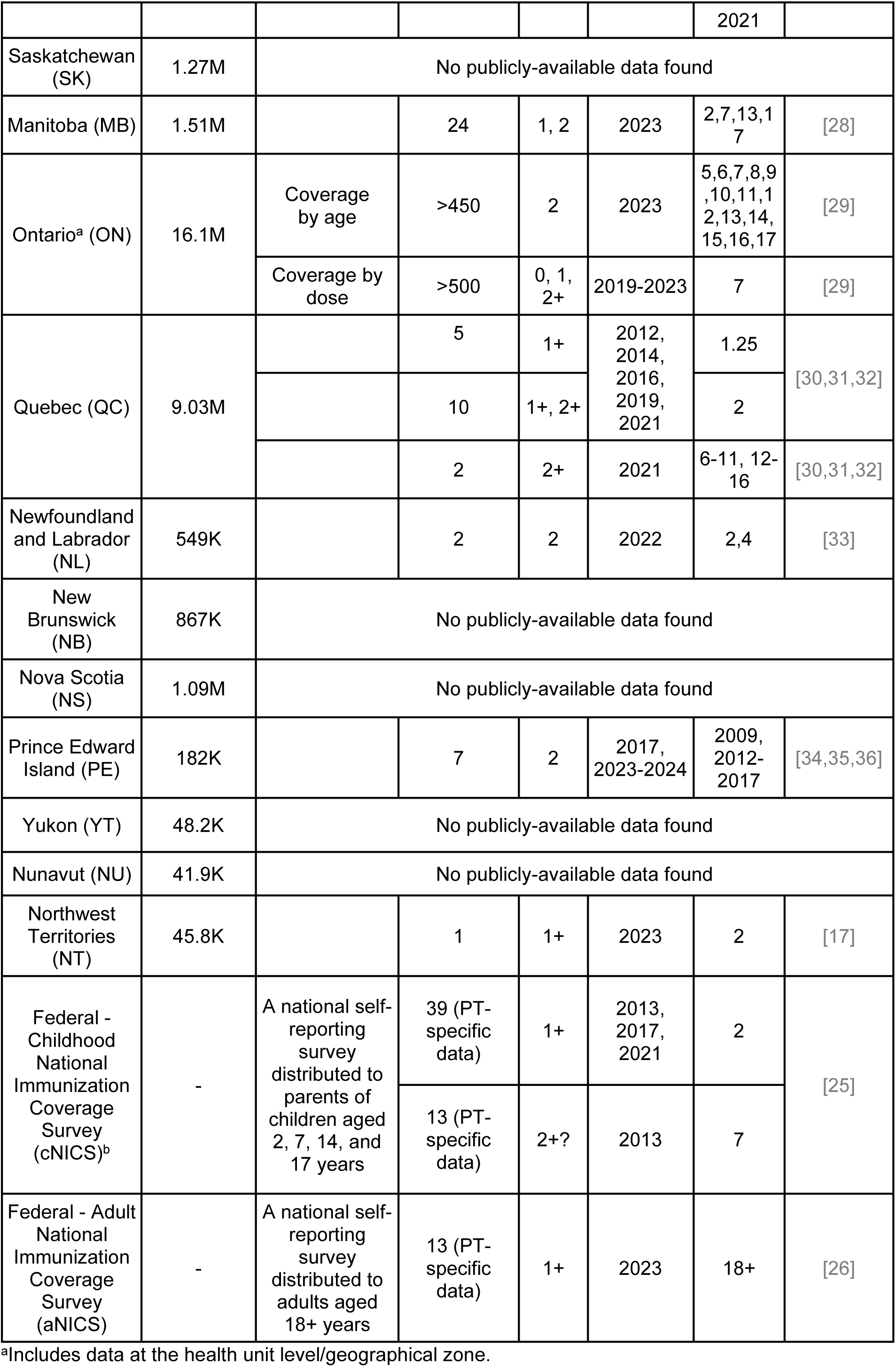
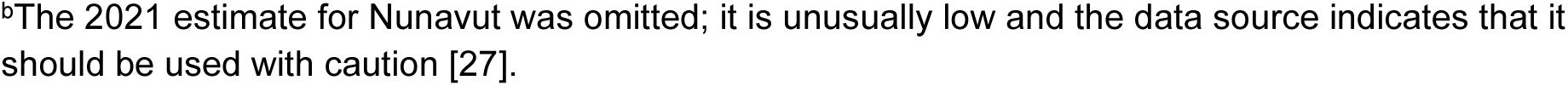
Summary of collated publicly-available vaccine coverage estimates. Data are taken from a range of PT and national-level sources [18,19,27–38].

| Region name<br>(Province or<br>Territory<br>Abbreviation) | Population<br>size<br>(2026) [39] | Description | Number of<br>available<br>vaccine<br>coverage<br>estimates | Dose<br>type | Years<br>reported | Age<br>cohorts<br>covered<br>(years)<br>or birth<br>years | Reference |
| --- | --- | --- | --- | --- | --- | --- | --- |
| British<br>Columbia (BC) | 5.66M |  | >450 | 1<br>2 | 2013-2023 | 2<br>7 | [27] |
| Alberta<sup>a</sup> (AB) | 5.05M |  | >10,500 | 1, 1+ | 2014-2023 | 1997- | [16] |
|  |  |  |  |  |  | 2021 |  |
| Saskatchewan (SK) | 1.27M | No publicly-available data found |  |  |  |  |  |
| Manitoba (MB) | 1.51M |  | 24 | 1, 2 | 2023 | 2,7,13,17 | [28] |
| Ontario<sup>a</sup> (ON) | 16.1M | Coverage by age | >450 | 2 | 2023 | 5,6,7,8,9,10,11,12,13,14,15,16,17 | [29] |
|  |  | Coverage by dose | >500 | 0, 1, 2+ | 2019-2023 | 7 | [29] |
| Quebec (QC) | 9.03M |  | 5 | 1+ | 2012, 2014, 2016, 2019, 2021 | 1.25 | [30,31,32] |
|  |  |  | 10 | 1+, 2+ |  | 2 |  |
|  |  |  | 2 | 2+ | 2021 | 6-11, 12-16 | [30,31,32] |
| Newfoundland and Labrador (NL) | 549K |  | 2 | 2 | 2022 | 2,4 | [33] |
| New Brunswick (NB) | 867K | No publicly-available data found |  |  |  |  |  |
| Nova Scotia (NS) | 1.09M | No publicly-available data found |  |  |  |  |  |
| Prince Edward Island (PE) | 182K |  | 7 | 2 | 2017, 2023-2024 | 2009, 2012-2017 | [34,35,36] |
| Yukon (YT) | 48.2K | No publicly-available data found |  |  |  |  |  |
| Nunavut (NU) | 41.9K | No publicly-available data found |  |  |  |  |  |
| Northwest Territories (NT) | 45.8K |  | 1 | 1+ | 2023 | 2 | [17] |
| Federal - Childhood National Immunization Coverage Survey (cNICS)<sup>b</sup> | - | A national self-reporting survey distributed to parents of children aged 2, 7, 14, and 17 years | 39 (PT-specific data) | 1+ | 2013, 2017, 2021 | 2 | [25] |
|  |  |  | 13 (PT-specific data) | 2+? | 2013 | 7 |  |
| Federal - Adult National Immunization Coverage Survey (aNICS) | - | A national self-reporting survey distributed to adults aged 18+ years | 13 (PT-specific data) | 1+ | 2023 | 18+ | [26] |

<sup>b</sup>The 2021 estimate for Nunavut was omitted; it is unusually low and the data source indicates that it should be used with caution [27].

#### Preparing Canadian model training data

We undertook a number of preprocessing steps to prepare the raw, collated data for input into our models. We excluded sub-PT-level estimates as our model was built to predict on the PT-level, resulting in approximately 200 data points (Table 1). Our data was then cleaned, including ageing estimates to present day (2026), deduplicating, and filtering down to *at least one (“1+”) dose* estimates for ages 5-21 year olds, as detailed in the Supplementary Material (Data preparation). In total, the refined dataset included 85 data points for at least one dose across the 13 PTs. PT-level sample sizes are listed in Table 2. Coverage estimates ranged from 68.0% to 98.0%, with a mean of 89.3% and a standard deviation of 5.2%. We also identified a further 58 data points for *two or more doses* - these are not used to train models but serve as an estimate of the lower bound for our model estimates of at least one dose coverage (green points in Results Figure 4).

**Table 2:** Number of vaccine coverage estimates by region in the filtered model-input dataset (1+ doses, current ages 5-21). Here, cNICS and aNICS estimates are merged with PT-specific data.

| BC | AB | SK | MB | ON | QC | NL | NB | NS | PE | YT | NU | NT |
| --- | --- | --- | --- | --- | --- | --- | --- | --- | --- | --- | --- | --- |
| 12 | 17 | 4 | 5 | 8 | 11 | 4 | 4 | 4 | 4 | 4 | 3 | 5 |

### 2.2 Model

#### Model formulation

We estimated vaccine coverage using a Gaussian Process model with two independent variables; current age and region:

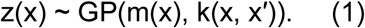

Here, z(x) is the vaccine coverage at each age-region pair x, m(x) is the expected coverage at x (a coordinate for age and a region metric), and k(x,x′) is a covariance function describing the strength of correlation between age-region pairs x and x′.

This model implies that the joint distribution of vaccine coverage estimates for each age-region combination is a multivariate normal distribution. We used a minimally informative prior with mean equal to the global sample mean across all PTs and ages: prior ∼ Normal(m, K_oo_)), for covariance matrix K_oo_ defined by the chosen covariance function and evaluated pairwise on all observed age-region combinations.

Conditioning on observed data, the posterior distribution was given analytically by posterior ∼ Normal(<u>μ</u>, Σ²), for

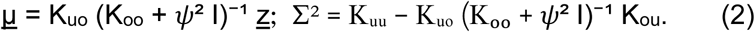

Here K_ij_ denotes the covariance matrix between observed (o) and/or unobserved (u) points (i,j ∈ {o, u}), *ψ* is the measurement error, I is the identity matrix, and z denotes the vector of observed vaccine coverages [39].

This GP formulation allows us to treat the covariates, age and region (metric), as continuous though in the data, age is measured discretely by year. Note that since vaccine coverage is bounded between 0 and 1, and Gaussian Processes typically assume the response variable can take on any real value, we modelled the *logit-transformed* vaccine coverage throughout and inverse-transform back before presenting results. Estimates were allowed to deviate by a normally-distributed variance *ψ*^2^ from data points (calculated in the logit space) to account for possible measurement error.

#### Covariance functions

Central to the Gaussian Process was the specification of a covariance function that quantifies similarity between inputs. We considered two commonly used covariance functions [40]:

Squared exponential:

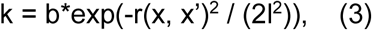

Exponential:

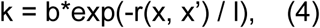

Here, r(x, x’) denotes the distance between age-region pairs x and x’, l is a lengthscale parameter, and b is a multiplicative factor. Our goal was to estimate the parameters l and b, since knowing these would allow us to infer vaccine coverage in age-region pairs where vaccine coverage is not reported. The squared exponential function assumes a gradual decay in correlation between nearby points, while the exponential function assumes a more rapid decay, implying weaker correlation over short distances.

#### Similarity indicators and determinants of vaccination

Our GP method leveraged relationships between observations based on covariates (age, region) to estimate unobserved values. This is encoded in the covariance functions described above, which rely on a distance metric between inputs, r(x, x’). In this paper, we treated age as a continuous variable and assumed that vaccine behaviours change gradually in the population, such that coverage at nearby ages is assumed to be similar. To relate regions in a similar way that is meaningful to our application, it was important to first identify determinants of vaccination.

Previous studies of vaccination of children in Canada have identified that socioeconomic inequalities, such as lower income, were associated with observed vaccination coverage heterogeneity [41–44]. The well established literature on health inequalities, i.e. systematic and socially produced, has shown how disadvantaged groups, for example people living in poverty, were more likely to suffer poorer health outcomes [45]. For example, the Gini coefficient is a measure of income inequality and has been found to be significantly associated with health inequality [46]. In conjunction with this, an examination of the structural determinants (governance, laws, policies, regulations, budgets and institutional practices), other social determinants (values, beliefs, culture, and norms), or factors within which individual-level behaviours take place, highlighted that numerous barriers exist and persist including mistrust of science and institutional mistrust [47]. This can further contribute to vaccine hesitancy and reduced rates of vaccination.

We hence chose to explore four indicators for relating Canadian PTs: *Gini index*, *the percentage of children in low-income families*, *the percentage of reported vaccine hesitancy among parents (pre-pandemic)*, and an alternate proxy for vaccine hesitancy, *the proportion of children who did not get vaccinated against COVID-19 (in 2022)*. Distances between PTs in our model were thus calculated using these socioeconomic and behavioural indicators for Euclidean distance r(x, x’) rather than geographic proximity. The *Gini index* provided a general measure of income inequality between PTs, while the other two indicators are more directly related to vaccination behaviours in Canada [41,42]. Gini index data was taken from Statistics Canada and was based on adjusted household after-tax income (mean of 2015 and 2020 data) - the coefficient ranges from 0 to 100%, where 0% indicates complete equality and 100% indicates complete inequality [20]. The *percentage of children in low-income families* data was obtained from the 2022 Health Inequalities Data Tool (Low-Income Cut Off, age-standardized rate calculated as 100*numerator/denominator) [48]. The *percentage of reported vaccine hesitancy among parents (pre-pandemic)* data was taken from the 2017 cNICS survey (refuse all + hesitant) [49]. Vaccine hesitancy data was not available for all PTs and was reported as a combined metric across PTs for the “Atlantic region” (NB, NL, NS, PE) and the “Northern region” (NT, NU, YT). Lastly, the *proportion of children who did not get vaccinated against COVID-19* data was taken from Health Infobase (ages 5-11), using data from 24th April 2022 when most PTs had achieved vaccine coverage saturation and before individuals aged out of the 5-11 cohort [50]. Indicator values by PT are reported in Supplementary Table S1. All indicators were ordered such that higher values correspond to expected decreases in healthcare-seeking behaviours.

#### Model hyperparameters

This model formulation yielded five hyperparameters (k, l1, l2, b, *ψ*). Broadly, these hyperparameters quantify how the model should leverage relationships between the covariate values in order to predict the response variable, and in particular, how much weight to put on “nearby” observations in performing inference of missing points. Weights are primarily defined by k, the covariance function, which depends on b, a multiplicative scaling factor, and “lengthscales” l1 and l2, which serve to define the distance between points in covariate space. Non-zero measurement error *ψ* softens the constraint that GP model predictions must pass precisely through observed values, which encodes the reality that observed measurements may not be perfect. Hyperparameters are catalogued in Table 3 with notes for further interpretation.

**Table 3:**
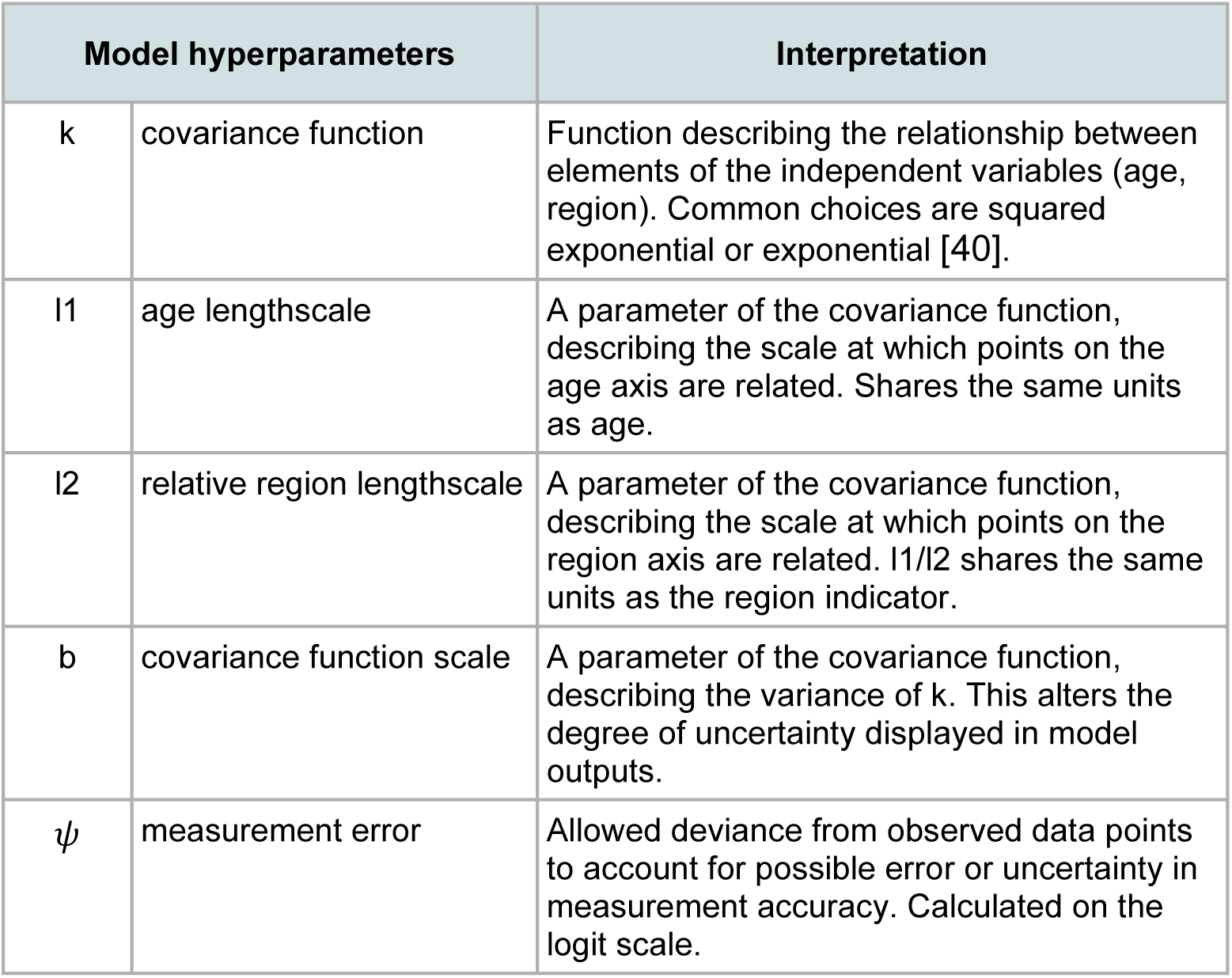
Hyperparameters of the Gaussian Process model.

The covariance functions defined above assumed a single lengthscale l. To accommodate two dimensions in our model, we assigned separate lengthscales to age and region. We rescaled the region direction by l2 then used one common lengthscale l1 across the whole model. l2 should hence be interpreted as the *relative multiplicative* region lengthscale (relative to l1), with l1 as the age lengthscale. Large values of l2 correspond to little to no relation between regions.

#### Model selection and validation

Model (hyperparameter) selection was performed using leave-one-out cross-validation (LOO-CV), with log posterior predictive density (LPPD) as the error metric at test points. This procedure was performed on an exhaustive grid of the four main hyperparameters (k, l1, l2, b), with each one restricted to a plausible search range (see Supplementary Material, *Details of model selection*, Range searched). The combination of plausible hyperparameter values with largest absolute LPPD, for fixed measurement error *ψ*, was chosen for each PT-relation (Table 4). Methodological details and full model selection results are included in the Supplementary Material.

**Table 4:** Optimal hyperparameter values selected for using leave-one-out cross-validation across the four PT relation indicators considered. l1 is in units of age (years). The column l1/l2 has been included to aid with interpretation, since its units match that of the relevant indicator (percentage); l2 is defined as the region lengthscale relative to l1, so it is more difficult to interpret directly. The maximum plausible l2 values considered (beyond which our 2D Gaussian Process model essentially collapses to an age-only model) were l2 = 50, 250, 8, and 5 for the Gini, Low Income, Vaccine Hesitancy, and COVID-19 Unvaccinated models respectively. Measurement error ψ is set to the value 0.05 and is not included in the hyperparameter optimisation. See the <u>Model selection and</u> <u>validation</u> methods subsection for more details.

| Indicator | Posterior parameter values used |  |  |  |  |  |
| --- | --- | --- | --- | --- | --- | --- |
| | k | l1 | l2 | l1/l2 | b | $\psi$ |
| Gini Index | Exponential | 2.5 | 25 | 0.1 | 0.5 | 0.05 |
| Low Income | Exponential | 2.5 | 250 | 0.01 | 0.5 | 0.05 |
| Vaccine Hesitancy | Exponential | 2.5 | 4.0 | 0.63 | 0.5 | 0.05 |
| COVID-19 Unvaccinated | Exponential | 2.5 | 3.5 | 0.71 | 0.5 | 0.05 |

For additional validation of our modelling approach, we also performed simulation-based validation on synthetic data sets and a second LOO-CV leaving out data for an entire PT each time to explore the model’s ability to predict vaccination rates at the PT level (LOOP- CV). Standard LOO-CV on individual observations may underestimate prediction error when nearby observations are autocorrelated, as the model can draw heavily on neighbouring points to predict a held-out one. The addition of the LOOP-CV analysis aims to combat this limitation, holding out all age observations for an entire PT simultaneously to assess whether the model can generalise to a completely unobserved region. See Supplementary Material for further details.

### 2.3 Model testing

#### Application to England’s data

We first tested our modelling framework against data for England, a country similar to Canada but with more complete vaccine coverage data, to further validate the reliability of our method and to quantify model accuracy. England was a natural comparator country to Canada as it has similar public health infrastructure (*i.e.*, free healthcare at the point of use and all essential routine vaccines offered free of charge), along with publicly-available data on vaccine coverage and demographic indicators by region. The nine regions of England have similar population sizes (ranging from approximately 2.8 million in the North East to 9.6 million in the South East) to some of the larger Canadian PTs (5.1 million for Alberta to 16.1 million for Ontario; see Table 1). Although England does not have smaller rural populations akin to the Canadian territories, variation in coverage is still observed across regions [51,52].

The vaccine coverage data used for England were for 2014-2025, and represent vaccination for 5-year olds with at least one measles dose (ages 6-17 in 2026). Data was publicly-available for all nine regions of England (e.g. London, South West, South East etc.) resulting in 108 data points in total. Coverage estimates ranged from 84.4% to 97.6%, with a mean of 94.4% and a standard deviation of 2.4%. All data was taken from the UK Health Security Agency data dashboard [52]. Relation of geographic areas (regions) used the Gini index for total wealth from ONS 2016-2018 (Supplementary Table S3) [53]. The Gini model framework was chosen as the primary testing framework as it performed best in assumptions tests in the Canada model (described below).

To test our model framework, we repeatedly subsampled the England data to the same sparsity as the Canadian data (in the overlapping age range of 6-17) to form a model training set; the rest of the data formed a test set. By “same sparsity”, we mean that, for each of the nine English regions, we randomly sample a number of observations (without replacement) from the number of model training points by PT in the Canadian data for ages 6-17, and then randomly sample that number of observations from the England data for the train set. For instance, in one replicate we may select up to two English regions to have a training sample size of 10 observations (sample sizes for BC and QC for ages 6-17), but only up to one with the minimum sample size of 2(NU). See Supplementary Methods, *Model testing - England comparison* for more details.

Within each experimental replicate (subsampling; N = 40), we used the training set to first select model hyperparameters (as described in the *Model selection and validation* subsection) and then to train the model. Finally, we computed an accuracy score at each left-out test point, which captured the probability that the fitted model’s predictions would come within an absolute 5% of the true (test) value. This measure allowed us to quantify model accuracy in a readily-interpretable way, compared to standard, but rather intractable, measures like the log posterior predictive density.

Due to the sparsity of the data in several PTs, the above sampling process can hold out large age segments of the England data. Having differences in sparsity preserved by region makes for a more difficult prediction problem than simply randomly subsampling the England data to the same overall number of observations as are included in the Canadian data (see Supplementary Methods, *Model testing - England comparison* and Supplementary Figure S3).

#### Test of similarity assumptions for model covariates

Like any regression model, a Gaussian Process model assumes relatedness between the independent variable(s), *i.e.* that ‘nearby’ ages (defined with lengthscale parameter l1) will have similar vaccination coverage and that ‘nearby’ regions will also have similar coverage (defined with lengthscale l1/l2). We tested these two assumptions to further investigate whether a GP model with these covariates could plausibly produce accurate predictions for the Canadian data.

We used a randomization (permutation) test to assess whether vaccine coverage varied smoothly with age, *i.e.*, whether ages that are closer together tend to have more similar coverage than expected by chance (assuming region is fixed). The null hypothesis was that vaccine coverage was not structured by age. The alternative hypothesis was that ages that are closer together have more similar vaccine coverage than expected by chance - coverage varies smoothly with age. For each age-gap group (1, ≤2, ≤3 years apart), the test statistic was the mean absolute pairwise coverage difference. Specifically, for each group g, we computed the average of |cᵢ − cⱼ| across all age pairs (i, j) in that group. The null distribution was generated by randomly shuffling coverage values across ages within each region (1000 iterations). This removed any real age structure while preserving the marginal distribution of coverage within each region. We computed a one-tailed p-value: the proportion of null replicates in which the mean pairwise difference was less than or equal to the observed value. A small p-value indicates that the observed differences between nearby ages were smaller than expected under randomness.

We repeated the same test for region similarity, performing it once per region similarity indicator (grouped by age). Since each indicator has a different range, we considered four relative similarity categories based on percentages of each indicator’s value range: ≤10%, ≤25%, ≤50% of range, and all pairs. For example, the Gini index spans 7.25 percentage points across PTs (28.25–35.50; Supplementary Table S1), so “within 10% of range” includes PT pairs whose Gini values differ by at most 0.725 percentage points, “within 25% of range” includes pairs differing by at most 1.8125 percentage points, and “within 50% of range” includes pairs differing by at most 3.625 percentage points.

## 3 Results

### 3.1 Testing model accuracy (England)

We first present results of testing our Gini modelling framework on subsamples of data from England, which has more complete data (Figure 2). Repeated subsamplings of England’s data show that the model can predict held-out test points with reasonable accuracy for data as sparse as Canada’s (mean accuracy 82% across test points from all 40 subsamplings).

**Figure 2:**
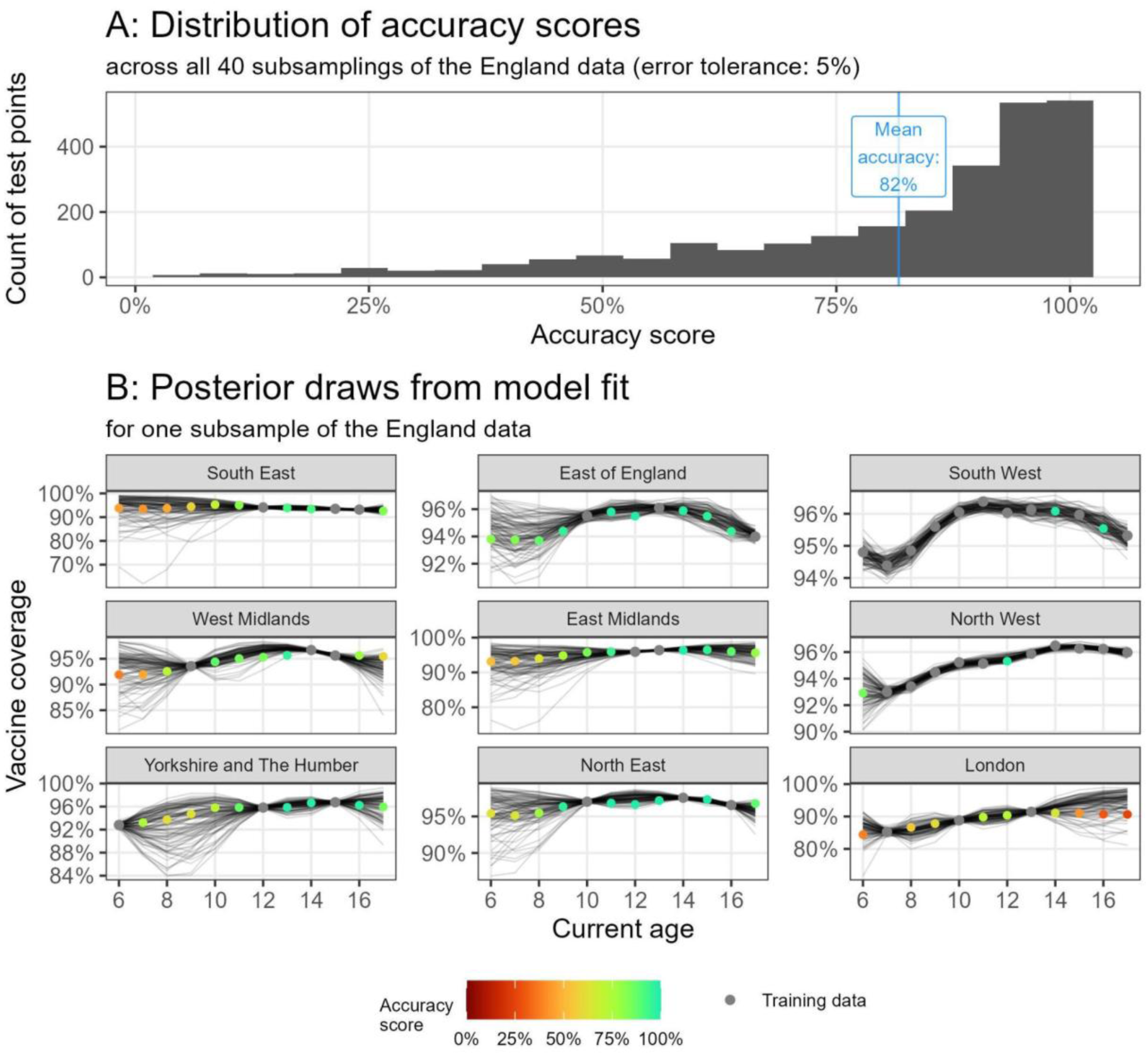
Model accuracy against held-out test points when applied to England data subsampled to match the sparsity of the Canada data. Accuracy was quantified as the probability that the fitted model’s predictions fall within 5% of each true (test) value. Hyperparameter optimization was performed anew for each subsampling. Panel A: aggregated distribution of accuracy scores (one score per test point) across all 40 subsamplings of the England data. The model achieved a reasonable mean accuracy (82%). Panel B: Model estimates (posterior draws; lines) from fitting the model to one subsampling (grey points), along with test data (points coloured by the corresponding accuracy score). Regions of England are shown as facets, ordered by their Gini index (ascending).

### 3.2 Testing age and region similarity assumptions (Canada)

Table 5 reports results from significance tests for the similarity in vaccine coverage using 1000 random permutations in the Canada model. We observed a significant relationship for coverages up to a moderate distance for age pairs (≤3 years) and between a near and moderate distance for region pairs when using the Gini Index. Figure 3 shows the distribution of absolute pairwise differences in vaccine coverage categorized by similarity level for the age and region (Gini) similarity assumptions underlying our GP model. The same plots for the Low Income, Vaccine Hesitancy, and COVID-19 Unvaccinated indicators are shown in the Supplementary Material (Supplementary Figures section).

**Figure 3:**
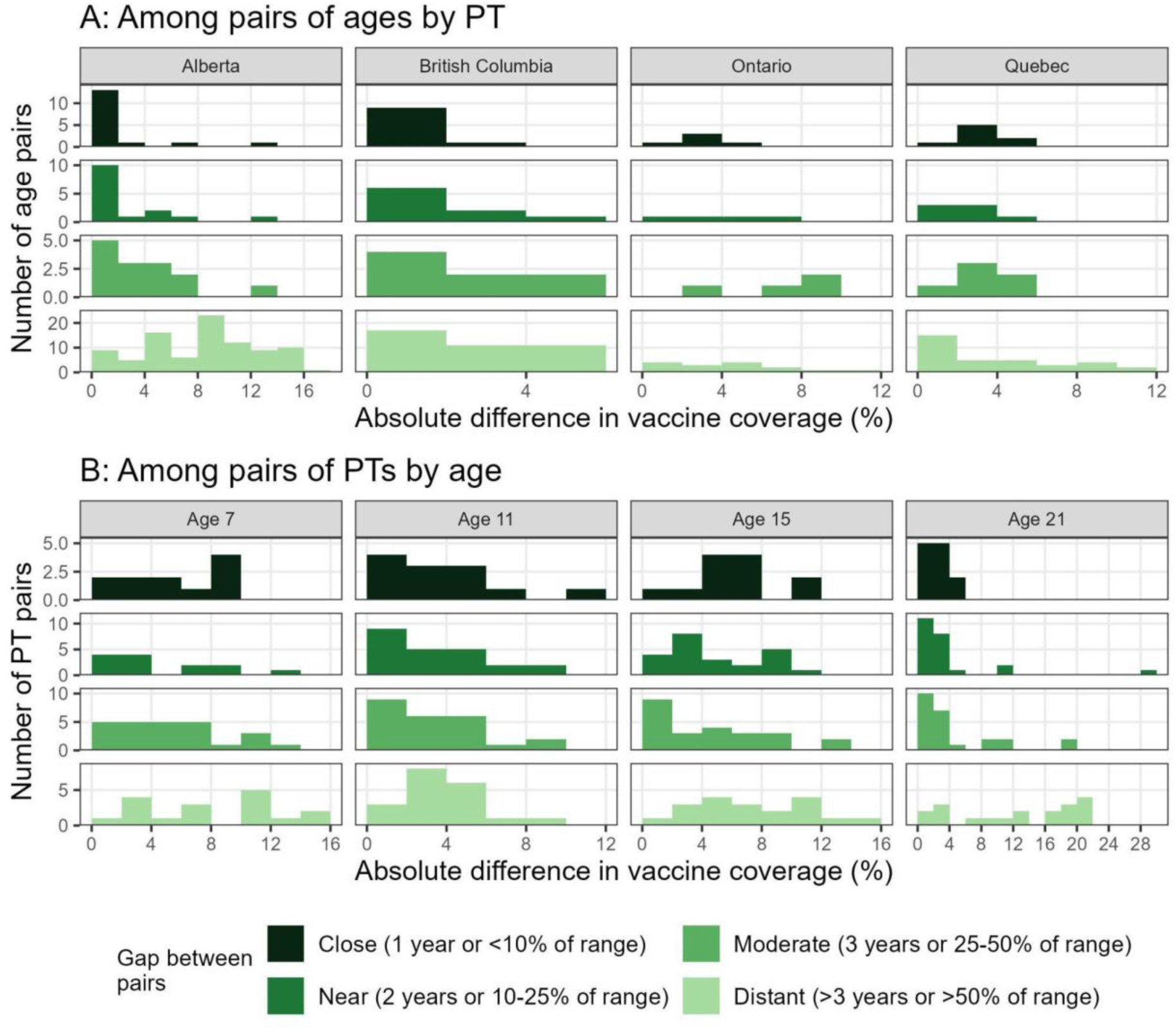
Visualizing the pairwise similarity in vaccine coverage based on the distance between corresponding covariates (age or Gini index). We show the distribution of absolute difference in vaccine coverage among pairs of ages for fixed PT (panel A) and among pairs of PTs for fixed age (panel B). Bars are coloured by the gap between covariate values, split into four categories: close (1 year for age or <10% of Gini index range for PT), near (2 years or 10-15% of range), moderate (3 years or 25-50% of range), and distant (>3 years or >50% of range). All indicator values are reported in Supplementary Table S1. We only show PTs and ages with coverage pairs across all gap categories. ‘Nearby’ ages (1-3 years apart) generally had lower absolute pairwise differences in coverage compared to those further (>3 years) apart. Regions at near to moderate distances from each other (10-50% of indicator’s range) tended to have lower absolute pairwise differences in coverage compared to those very close together (<10% of range) or those very far (>50% of range).

**Table 5:** Statistical significance of similarity in vaccine coverage depending on the distance between covariate measures (age and region indicators). Tests used nested distance thresholds: within close range (≤1 year or ≤10% region indicator range), within near range (≤2 years or ≤25% indicator range), within moderate range (≤3 years or ≤50% indicator range), or all pairs. For each nearness category, we used a permutation test to estimate how likely it is, under random chance, to observe a mean absolute pairwise difference in vaccine coverage as small as the one observed in the true data (one-sided p-value). Smaller p-values indicate that ‘nearby’ observations have more similar coverage than expected assuming there is no relationship in vaccine coverage for nearby ages and regions. Significant relationships were found for ages up to a moderate distance and for regions within near and moderate distance based on the Gini index.

| Covariate | Indicator | p-values |  |  |
| --- | --- | --- | --- | --- |
|  |  | Within **close** range<br>(≤1 year or ≤10% indicator range) | Within **near** range<br>(≤2 years or ≤25% indicator range) | Within **moderate** range (≤3 years or ≤50% indicator range) |
| Age | Age | <0.001 | <0.001 | <0.001 |
| Region | Gini Index | 0.203 | 0.001 | <0.001 |
|  | Low Income | 0.419 | 0.510 | 0.485 |
|  | Vaccine Hesitancy | 0.375 | 0.383 | 0.041 |
|  | COVID-19 Unvaccinated | 0.011 | 0.003 | 0.081 |

### 3.3 Model estimates of vaccine coverage (Canada)

After model testing, our main result was the application of GP models to estimate vaccine coverage in Canada (Figure 4). Optimal hyperparameters from model selection are reported in Table 4. For the Gini Index and Vaccine Hesitancy, information on vaccine coverage appears to be shared between PTs (see table; l1/l2 greater than smallest gap between indicator values, but less than overall indicator range). For the Low Income model, little information is shared between PTs (l1/l2 equal to the smallest gap between indicator values), effectively reducing the model to age-only, which suggests this indicator did not carry much predictive power when relating regions in a GP model with age as the second covariate.

**Figure 4:**
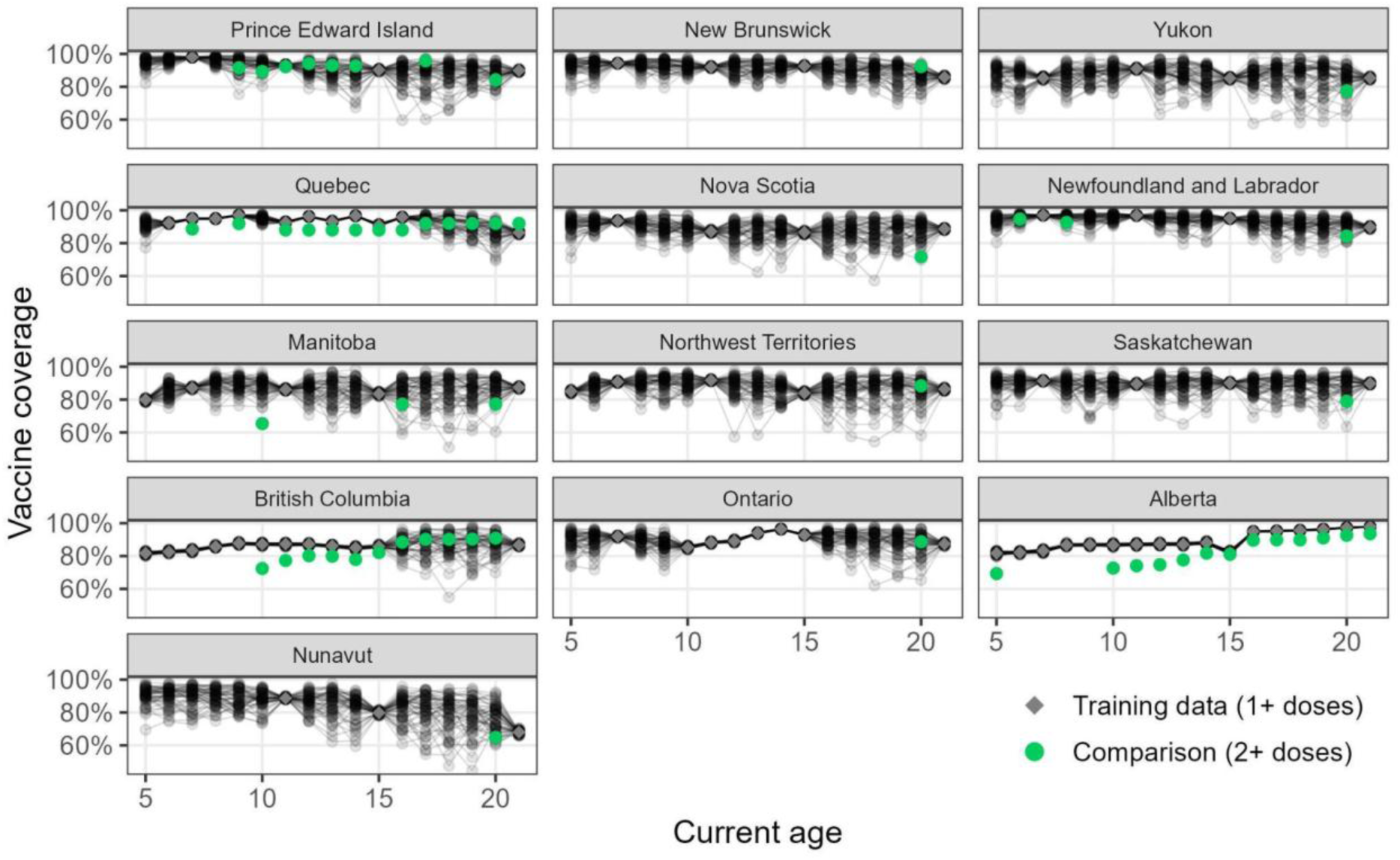
Model results for Canada showing estimates of vaccine coverage for at least one dose of a measles vaccine by age and province/territory (PT) for the Gini model. Black curves represent model estimates (posterior draws; 50 random draws). Grey points denote training data (1+ dose coverage), while green points show 2+ dose coverage as a comparison (not used to fit the model). PTs are shown as facets, ordered by their Gini index (ascending). Model estimates are provided with a moderate-to-high degree of uncertainty, with estimates in smaller age gaps between observations typically associated with lower uncertainty. The comparison to 2+ dose data suggests plausible 1+ dose estimates, as we expect at least two dose coverage to be smaller than that of at least one dose.

In exploring further (Supplementary Figure S2), we observed that for our Gini and Vaccine Hesitancy modelling frameworks, the full (age and region) model was preferred to a proxy age-only model. In particular, estimates for Manitoba and Northwest Territories benefitted greatly from the additional PT dimension when using the Gini Index. This result further supports the use of the Gini Index and Vaccine Hesitancy indicators to relate regions in inferring vaccine coverage jointly with age.

Model estimates from the Gini index framework are shown in Figure 4. The same figures for the other three PT relation indicators are included in the Supplementary Material (Supplementary Figures section). We also compared our model estimates to unseen data, namely 2+ dose data described in Table 1. This is overlaid in green for comparison in Figure 4. Our model inferences appear plausible but there is a clear need for better data from harmonised data collection and methodology, in order to obtain meaningful model estimates.

## 4 Discussion

In this work, we have developed proof-of-concept methodology for estimating missing vaccine coverage data, motivated by the Canadian setting, which is currently undergoing a multi-year measles outbreak. Our approach focuses on relating age cohorts and regions to make the most of the sparse publicly-available data. Overall, this was a challenging imputation problem, with many missing data points to be inferred. For example, when using the Gini Index framework, 85 data points were available for a total of 221 age-region pairs (38%), meaning 62% of data was missing and had to be inferred. For the Vaccine Hesitancy framework (Supplementary Material), the aggregation of data from Northern PTs and Atlantic PTs reduced the proportion of missing data to 51.5%, but at the expense of generalizing coverage across entire regions with multiple public health authorities. Due to the small number of usable, publicly-available data points in Canada (85), we focused on methodology and quantifying uncertainty rather than exact prediction. We developed a flexible Gaussian Process modelling framework to identify strengths and weaknesses of the approach.

We applied our methodology to subsamplings of England’s very complete data, mimicking the sparsity of Canada’s data, in order to validate our approach by computing accuracy scores based on held-out true values (Figure 2). This was done for the Gini modelling framework to understand if the GP approach was in principle a plausible methodology. While the model generated estimates with relatively high (82%) accuracy for England’s data, these input data varied less than Canada’s data (sample standard deviation of 5.2% for Canada, 2.4% for England) and so may represent an easier prediction problem. This difference may, in part, be due to the fact that England’s healthcare system is centrally-run through the National Health Service, while Canada’s is federated through individual provinces and territories; the latter means that vaccine coverage estimates are collated across multiple sources, which could lead to higher variation in the data. However, there may also be other population differences between the two countries that incur different levels of variance.

We continued to investigate the plausibility of our approach by testing key GP model assumptions; namely, that vaccine coverage should be similar for ‘similar’ ages and regions (Table 5 and Figure 3). Distance between regions was quantified using four plausible indicators for relating vaccination uptake behaviour among PTs: the Gini index (which captured income inequality), the percentage of children in low-income families, the percentage of parents who report being vaccine hesitant (pre-pandemic), and as a proxy for more recent vaccine hesitancy, the percentage of children unvaccinated against COVID-19 in 2022. The choice of indicator was fundamental to our underlying GP structure. Our testing found statistically significant relationships for age gaps 3 years and under and regions at intermediate distance as quantified by the Gini index. These tests broadly supported the relation of age cohorts and regions in inferring missing vaccine coverages and suggested that some indicators relating regions may be more predictive than others when using the GP model approach. However, this evidence was limited. Permutation tests were performed for one covariate at a time, conditional on the other being fixed, whereas GPs are fit jointly across covariates. Significant results from the permutation tests hence do not guarantee that GP models applied to this data would accurately predict missing observations.

Finally, we applied GP models to the Canadian data to provide estimates of coverage, focusing on the Gini index as our main indicator. When there was a small age gap between observed points, the model inferred the missing point(s) with moderate uncertainty (Figure 4, *e.g.*, age 10 in Quebec). Larger age gaps yielded higher uncertainty, but the uncertainty is quantified, which can be useful for input into disease models (*e.g.*, when performing a sensitivity analysis).

Our work motivates the need for more standardized data sharing in Canada, including the release of vaccine coverage estimates as numbers of individuals so that careful adjustments can be made to age data to present day. Our proof-of-concept also further motivates investigation into how best to utilise different indicators in combination.

### The need for statistical modelling

Estimating vaccine uptake across geographic regions was non-trivial - uptake is shaped by many factors simultaneously, including socioeconomic conditions, political context, health system access, and cultural attitudes toward vaccination, without any single predictor dominating. The relationship between these factors and uptake are likely nonlinear and may vary in magnitude and even direction across countries, contexts, and time, making it difficult to specify a fixed functional form *a priori*. This issue is compounded by a limited mechanistic understanding of how these predictors translate into individual vaccination decisions.

Together, these features argue against using simple parametric regression models, which would require strong assumptions about the form of predictor-outcome relationships, and against approaches that pool data across countries.

A Gaussian Process (GP) model is well-suited to this problem. GPs generalize the multivariate normal distribution to function spaces (i.e. regression problems), allowing us to learn relationships between predictors and vaccine uptake directly from data within a given context without committing to a particular functional form [40]. Unlike black-box machine learning approaches, GP models are fully embedded in the Bayesian framework, allowing statistical information to be shared across inferred parameters, and giving access to priors, likelihoods, and posteriors, enabling principled uncertainty quantification and model selection. Their hyperparameters, such as the lengthscales of the covariance between observations, retain interpretable meaning and can be compared across settings, providing some insight into the structure of the learned relationships. GPs are also extensible: incorporating data from a new region or re-aggregating to a different geographic scale requires no fundamental change to the model. This combination of flexibility, interpretability, and statistical rigour made GPs a natural tool for interpolating levels of vaccine uptake.

### Determinants of measles vaccination in Canada

As noted in the Methods section, our GP modelling frameworks utilised data on possible determinants of measles vaccination in Canada to relate PTs in the absence of comprehensive vaccination coverage data. We considered a variety of metrics and selected four to be used with the GP methodology. While a multitude of factors may influence vaccination uptake in Canada, vaccine hesitancy has been identified as an important driver of undervaccination [54]. Vaccine hesitancy is a complex issue that has been on the rise globally and in Canada for the last several decades [55]. While parental socioeconomic status, education, and misinformation have been highlighted among multiple factors contributing to vaccine hesitancy internationally, these do not necessarily translate into direct determinants of vaccination [56,57]. Furthermore, unvaccinated groups are far from homogenous in their motivations. Studies in Canada found that some of these factors interact with differential effects across age groups with parental concerns about side effects and cultural mistrust repeatedly being raised [58–60]. Religious concerns and accessibility also stood out in Canada [47,61,62]. A further complication is the challenge of measuring the constructs related to vaccine hesitancy with measurement tools requiring psychometric validation in the populations under study. The non-direct relationship between potential factors and vaccination may explain in part why our results, while promising, struggle to paint the full picture of vaccination coverage. Indeed, a recent multi-country study of inequity and vaccination highlighted the need for a multivariate approach to better capture vaccination coverage [63]. In future work, we hope to explore further indicators in combination, to further strengthen modelling frameworks until better data becomes available.

### Limitations of our work

The Canadian data was of very limited quality: some PTs had very few data points available, while the cNICS/aNICS data was from self-reported surveys that could include reporting biases. Since we fit our model to data from numerous different sources, inconsistencies in data methodology could introduce unknown error and bias. There were also no data points for present ages 1-4 years in part due to delays in data releases, and so we could not produce reliable estimates for these epidemiologically-critical ages.

The model also used naive ageing of older estimates and hence did not account for changes within a birth cohort over time; catch-up vaccinations, emigration/immigration between PTs and in and out of Canada, and political and cultural changes over time were not captured.

For example, our primary vaccine hesitancy indicator used data from a 2017 survey and hence would not account for any changes in vaccine behaviours post-pandemic that are thought to have occurred in some PTs [12,64]. It is also worth noting that our GP model used a global prior mean which may skew results in age-region pairs; a point for future work is to include region-specific prior means. A final methodological limitation was that region indicator values must be unique - if any two PTs have the same indicator values, our GP model breaks down. For example, the Health Inequalities Data Tool summary data is reported to 1 decimal place but at this level of accuracy, some PTs had identical values for % of children in low-income families. We hence had to calculate this metric to 2 decimal places using a more-detailed downloadable dataset to produce unique indicator values for each PT [48]. For all of these reasons, our modelling frameworks are presented as a proof-of-concept and we do not expect our models to be used for exact predictions at this stage.

## Conclusion

Our work has highlighted the need for stronger, standardized, and publicly-available measles vaccine coverage data by age and PT in Canada. Our proof-of-concept Gaussian Process model utilised similarity between age cohorts and regions, and demonstrated a relatively high degree of accuracy when applied to subsampled data for England. The emerging STARVAX initiative is promising, but at time of writing, nine of Canada’s 13 provinces and territories are enrolled and PT-stratified data is not available [12,13]. Statistical models, such as the one described in this manuscript, can be useful in estimating coverage with quantified uncertainty, but sufficient data are needed to be able to make trustworthy predictions.

## Author contributions

All authors posed the research question and contributed to the design of this study. IP and RMM curated the publicly-available vaccine coverage dataset. RW developed the PT relation indicators. LH developed the modelling frameworks, with support from IP and SMK. LH and IP implemented simulation code and performed the analysis. All authors contributed to first writing and critical feedback of the manuscript and approved the final version for publication.

## Supporting information

Supplementary Material (revised)

## Acknowledgements

We wish to thank David Champredon for informative discussions on model evaluation techniques. We acknowledge the use of AI tools in coding assistance and in drafting and editing portions of the manuscript text. All AI outputs were reviewed and edited by at least one author.

## Data availability

All data and model code used in this article are available on GitHub at https://github.com/phac-modelling-hub/immunity-landscape.

