## Supplementary Material (revised) for "Making inferences with incomplete epidemiological data: a proof-of-concept estimating measles vaccine coverage across Canada"

#### Supplementary methods

##### Data preparation

###### Preparing Canadian model training data

As vaccine coverage data was pulled from multiple sources, there were a number of nuances that needed to be considered in the pre-processing stage, such as harmonising definitions of 1 or 2 doses and 1+ or 2+ doses. Additional pre-processing steps to produce the final model training data summarized in main text Table 2 were as follows:

*Data for a group of ages*

Where there was an estimate for a group of ages (e.g. 6-11), that coverage estimate was repeated for each age within the group.

*Ageing estimates to present day*

Our goal was to quantify the present-day immunity landscape. Therefore, all birth cohort data were aged to present day (2026). We did this by taking the age at estimate and adding the number of years that have elapsed between the production of the estimate (reporting year) and the present year (2026). This simplistic approach did not account for catch-up immunizations or population migration that may have occurred after these estimates were reported. Even with data on these processes, it would be difficult to adjust vaccine coverage *proportions* to account for these processes without knowing the denominators used to calculate these estimates.

*Different dose types*

Our model was not set up to predict more than one dose type. We therefore chose to produce results for coverage of *at least one dose* as it encompasses the largest subset of the original data (coverage reported for “1” and “1+” doses).

*De-duplicate estimates by age*

In several instances, there were multiple estimates reported for a present-day age cohort. In cases where there were duplicate estimates because they were reported both directly by provinces/territories (PTs) and from one of the national surveys (cNICS/ aNICS, the childhood and adult National Immunization Coverage Surveys respectively), we prioritized PT-reported data. If there were duplicates within the PT-reported data, we took the most recent estimate for that age cohort.

*Filtering the age range*

We restricted attention to individuals with a current age between 5 and 21 years. Age 5 was the first current age for which we had coverage data. We ended our present day age range at 21 to include the latest estimate from aNICS as an input (18+ in 2023).

In total, the refined dataset included 85 data points for *at least one dose* across the 13 PTs, after ageing to present day and filtering as described above (see main text Table 2 and grey points in Results figures, *e.g.*, Figure 4).

##### Considering bias between data types

Since the Canadian vaccine coverage data described above encapsulates multiple data types, we carried out supplementary analysis to explore possible biases. This analysis is available in our GitHub project repository, at: https://github.com/phac-modelling-hub/immunity-landscape/blob/main/notes/data-source-bias.qmd. We considered whether there may be evidence of bias between the national survey (aNICS/cNICS) and PT-reported vaccine coverage data for 1+ doses, where both were available for the same PT and current age. Results indicated little evidence of bias in the available survey data, when compared to PT-reported data.

#### Similarity indicators

As described in the main text (Methods, *Similarity indicators and determinants of vaccination*), our Gaussian Process model relied on a distance metric for PT. We chose to explore four indicators for relating Canadian PTs: *Gini index*, *the percentage of children in low-income families*, *the percentage of reported vaccine hesitancy among parents* (pre-pandemic), and *the percentage of children unvaccinated against COVID-19* (a 2022 vaccine hesitancy proxy). Values used are shown in Supplementary Table S1.

***Supplementary Table S1:*** ***PT relation indicators: values used.*** *See main text for full description. These values were multiplied by l2 in the model code. Data taken from StatCan, Health Inequalities Data Tool, cNICS, and Health Infobase* [1–4]. *NB: For the vaccine hesitancy indicator, data was combined for Atlantic region = NB, NS, NL, PE; and for Northern region = YT, NT, NU.*

| **Province or territory (PT)** | **PT relation indicator** | | | |
|---|---|---|---|---|
| | **% Gini index on adjusted household after-tax income** | **% of children in low-income families** | **% of parents who are vaccine hesitant** | **% of children unvaccinated against COVID-19** |
| British Columbia (BC) | 32.45 | 5.35 | 20.1 | 45.8 |
| Alberta (AB) | 33.90 | 4.66 | 16.3 | 54.3 |
| Saskatchewan (SK) | 31.20 | 5.13 | 18.1 | 43.3 |
| Manitoba (MB) | 30.90 | 7.20 | 19.1 | 41.4 |
| Ontario (ON) | 32.60 | 4.68 | 17.3 | 45.3 |
| Quebec (QC) | 29.35 | 3.73 | 26.9 | 37.8 |
| Newfoundland and Labrador (NL) | 30.65 | 3.16 | 17.0 (“Atlantic region”) | 12.3 |
| New Brunswick (NB) | 28.50 | 3.84 | 17.0 (“Atlantic region”) | 42.0 |
| Nova Scotia (NS) | 29.80 | 4.71 | 17.0 (“Atlantic region”) | 33.1 |
| Prince Edward Island (PE) | 28.25 | 3.76 | 17.0 (“Atlantic region”) | 32.1 |
| Yukon (YT) | 28.85 | 2.58 | 23.7 (“Northern region”) | 40.4 |
| Nunavut (NU) | 35.50 | 3.77 | 23.7 (“Northern region”) | 27.7 |
| Northwest Territories (NT) | 30.95 | 1.61 | 23.7 (“Northern region”) | 36.9 |

#### Model selection

Model equations for our Gaussian Process model are defined in the main text, with five unknown hyperparameters (Section 2.2 and Table 3). To identify the most suitable values, we performed hyperparameter selection/ model selection as described below. Here we are interested in how the model performs at observed points, which are partitioned into ‘training’ and ‘test’ points for the model selection process.

##### Derivation

We performed leave-one-out cross-validation (LOO-CV), using log posterior predictive density (LPPD) as the error metric, to identify suitable values for the hyperparameters in our model. The posterior predictive distribution can be calculated analytically as follows:

Let $\underline{x}$ and $\underline{z}$ be the training points (covariate coordinate with age and region metric) and training values (vaccine coverage) respectively, both vectors of length 85 (the total number of data points in our Canada model training dataset). The posterior predictive distribution with measurement error $\psi$ at a single test point $x_a$ is:

$$\mathcal{P}(z_a|x_a, \underline{x}, \underline{z}) \sim \mathcal{N}(\mu_a, \sigma_a^2)$$

where

$$\mu_a = \underline{k}(x_a, \underline{x})^T (K + \psi^2 I)^{-1} \underline{z},$$
$$\sigma_a^2 = k(x_a, x_a) - \underline{k}(x_a, \underline{x})^T (K + \psi^2 I)^{-1} \underline{k}(x_a, \underline{x}).$$

Here, $K$ is the 85x85 covariance matrix of training data points $(K_{ij} = k(x_i, x_j))$.

One can then evaluate the posterior predictive distribution at $z_a$ to produce the posterior predictive density at the test value, as:

$$\mathcal{P}(z_a|x_a, \underline{x}, \underline{z}) = \frac{1}{\sqrt{2\pi\sigma_a^2}} exp\left(-\frac{(z_a - \mu_a)^2}{2\sigma_a^2}\right).$$

Finally, taking logs gives:

$$lppd|_{z_a} = -\frac{1}{2} log(\sigma_a^2) - \frac{(z_a - \mu_a)^2}{2\sigma_a^2} - \frac{1}{2} log(2\pi).$$

For a fuller description, see Rasmussen & Williams [5]. Using a key simplification from the literature (e.g. section 5.4.2 in [5], section 2.2 in [6]), one can then calculate $\mu_a$ and $\sigma_a^2$ as:

$$\mu_a = z_i - \frac{[(K+\psi^2 I)^{-1} \underline{z}]_i}{[(K+\psi^2 I)^{-1}]_{ii}},$$
$$\sigma_a^2 = \frac{1}{[(K+\psi^2 I)^{-1}]_{ii}}.$$

where $K$ is now the full covariance matrix of observed (training+test) points, i is the index of the test (i=a), $\psi$ is the measurement error, $[A]_{ii}$ denotes the (i,i)-th element of a matrix A, and $[Ab]_i$ denotes the ith element of a vector $Ab$. Note that in our code, $K$ was simply $K_{oo}$ when running the full model. Hence the exercise of calculating LPPD with noise was largely reduced to extracting $(K + \psi^2 I)^{-1}$ from the full model [5,6].

##### Range searched

As noted in the main text, the above LOO-CV method was performed across the four main hyperparameters (k, l1, l2, b). Search ranges were as follows: k ∈ {squared exponential, exponential}; l1 ∈ {1, 1.25, 1.5, 1.75, 2, 2.25, 2.5}; l2 ∈ {0.1, 0.2, 0.5, 1, 1.5, 2, 2.5, 3, 3.5, 4, 6, 8, 10, 25, 50, 75, 100, 150, 200, 250, 500}. Additional sensitivity analyses explored b ∈ {0.5, 1.0, 1.5} (default value is 1). The l1 search range was informed by main text Figure 3, while for the l2 search range, we derived an *approximate*[1] meaningful upper and lower limit:

[1] NB: Strictly speaking, as we have a probabilistic model and exponential/squared exponential covariance functions, all equations in this derivation should be interpreted as approximations/ plausible bounds not exact.

Let $y_i$ denote the relational values assigned to each province, as defined in Supplementary Table S1. For example, for the Gini model, $y_1$ = 28.25 (Prince Edward Island), $y_2$ = 28.50 (New Brunswick), …, and $y_{13}$ = 35.50 (Nunavut). Recall l1 is the age lengthscale and l2 is defined as the *relative* multiplicative lengthscale of PTs to age. Hence, we can say that all PTs are related to each other (to some degree) on average if l1/l2 is approximately greater than the total range of y values, i.e. if: $\frac{l1}{l2} > y_n - y_1$.
Equivalently, there is little to no relation between any two regions on average if l1/l2 is less than the smallest difference in y values, i.e. if: $\frac{l1}{l2} < min(y_{i+1} - y_i)$.
Assuming some relation between at least some regions, we hence desire: $\frac{l1}{l2} \leq (y_n - y_1)$ and $\frac{l1}{l2} \geq min(y_{i+1} - y_i)$. Rearranging gives an approximate plausible range for l2 as:

$$\frac{l1}{y_n - y_1} \leq l2 \leq \frac{l1}{min(y_{i+1} - y_i)}.$$

For l2, we hence considered the following conservatively wide search ranges:

Gini model: 0.1 - 50,
Low Income model: 0.2 - 250,
Vaccine Hesitancy model: 0.1 - 8,
COVID-19 Unvaccinated model: 0.02 - 5.

All searches for l2 ran from 0.1 to 500 (CU: 0.02 to 500) but the 'best' hyperparameters chosen were restricted to the above plausible ranges. Choosing a larger l2 did not significantly affect model behaviour (see *Results of model selection* below).

Finally, $\psi^2$ was chosen to be smaller than elements of $K_{oo}$, on the logit scale, so as to represent the order of an error term and not dominate model equations. (For interest, in the Gini model, max($K_{oo}$) = 0.50, mean($K_{oo}$) = 0.15, median($K_{oo}$) = 0.06, and min($K_{oo}$) = 4.9e-10). We used $\psi$ = 0.05 throughout, i.e. $\psi^2$ = 0.0025.

#### Results of model selection

Results for best hyperparameter values for each modelling framework are shown in Table 4 in the main text. In Supplementary Figure S1 below we show the full results of LOO-CV. Note that b was set to its default value of 1 in this plot to illustrate, as varying b only affected model uncertainty not shape/behaviour.

***Supplementary Figure S1:*** ***Log Posterior Predictive Density (LPPD) heatmaps from model selection, for each modelling framework (Gini = Gini index model; LI = Low Income model; VH = Vaccine Hesitancy model; CU = COVID-19 Unvaccinated model).*** *Plots show l2 against l1, with varying measurement error $\psi$ shown as facets. Rows depict each modelling framework with k=exp (exponential) and sqexp (squared exponential). Axes are discrete categorical, not continuous. White regions indicate parameter combinations which performed poorly, with lppd ≤ -100. Note b=1 here.*

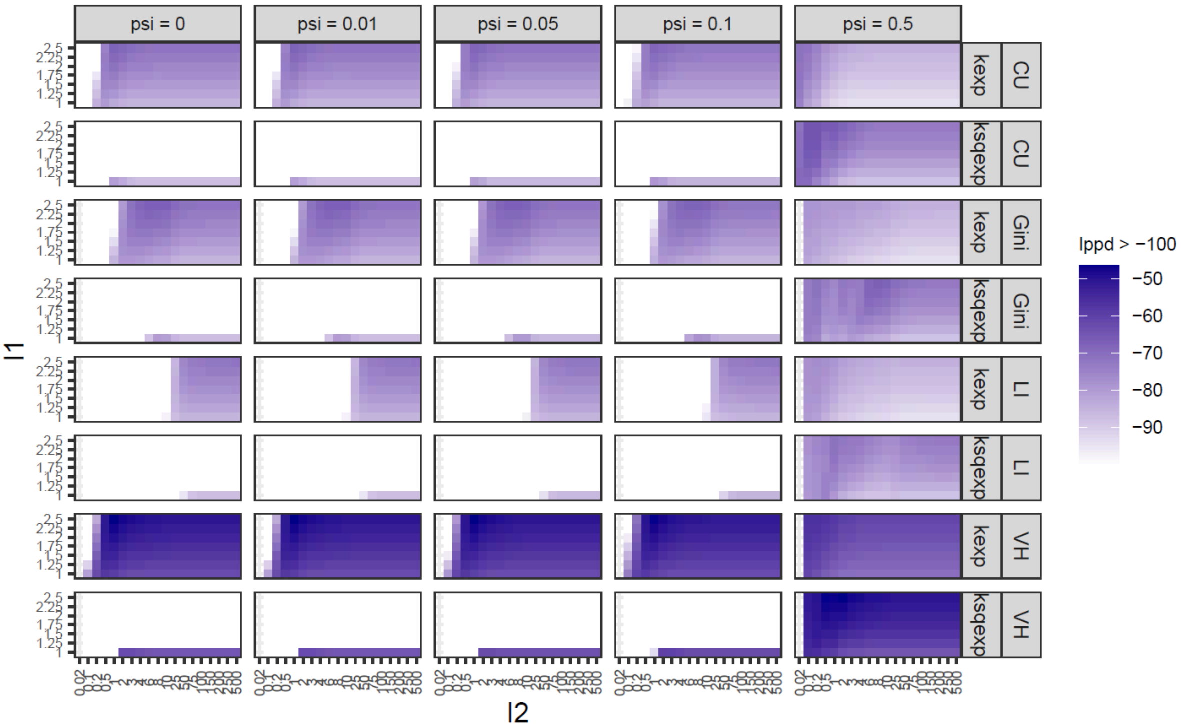


Our Low Income model favoured larger l2 (i.e. low relation between PTs), but increasing l2 beyond the meaningful plausible limits derived above (Gini: ~50; LI: ~250; VH: ~8; CU: ~5) made little difference to model selection or model behaviour. The 'best' hyperparameters chosen were hence restricted to the plausible ranges.

### Model validation

#### Leave-one-out by province/territory (LOOP-CV)

Leave-one-out cross-validation was also performed by leaving out all the data points for a single PT in turn, instead of one single data point at a time. We refer to this alternative analysis as leave-one-out by province/territory (LOOP-CV). Again we used LPPD as the error metric. This further validation step aimed to understand the level of PT predictability: some of our models favoured large l2 which could indicate that there was little relation between PTs or that there was little *extra relation* beyond the information given by the age lengthscale. It could be that the age groups of the data are sufficiently informative. To explore this, we compared LPPD per PT between the full model (best hyperparameters) and an age-only model. This is shown in Supplementary Figure S2. Here we were interested in how each PT compares to being left out of the full age+PT model, compared to just an age model. As a proxy, the "age-only" model was the full (age+PT) model but with very large l2 = 500.

***<u>Supplementary Figure S2:</u> Four plots comparing LPPD for each PT from the best full model to "age-only" proxy model.*** *Each panel shows each of the four modelling frameworks - (a) Gini, (b) Low Income, (c) Vaccine Hesitancy, (d) COVID-19 Unvaccinated. In each case, the difference in mean LPPD per PT between the full model and age-only model is shown. Hyperparameter values were as follows: For the Gini model framework, k=exponential, l1=2.5, b=0.5, psi=0.05, and l2 = 25 (best) and 500 (age). For the Low Income model framework, k=exponential, l1=2.5, b=0.5, psi=0.05, and l2 = 250 (best) and 500 (age). For the Vaccine Hesitancy model framework, k=exponential, l1=2.5, b=0.5, psi=0.05, and l2 = 4 (best) and 500 (age). For the COVID-19 Unvaccinated model framework, k=exponential, l1=2.5, b=0.5, psi=0.05, and l2 = 3.5 (best) and 500 (age).*

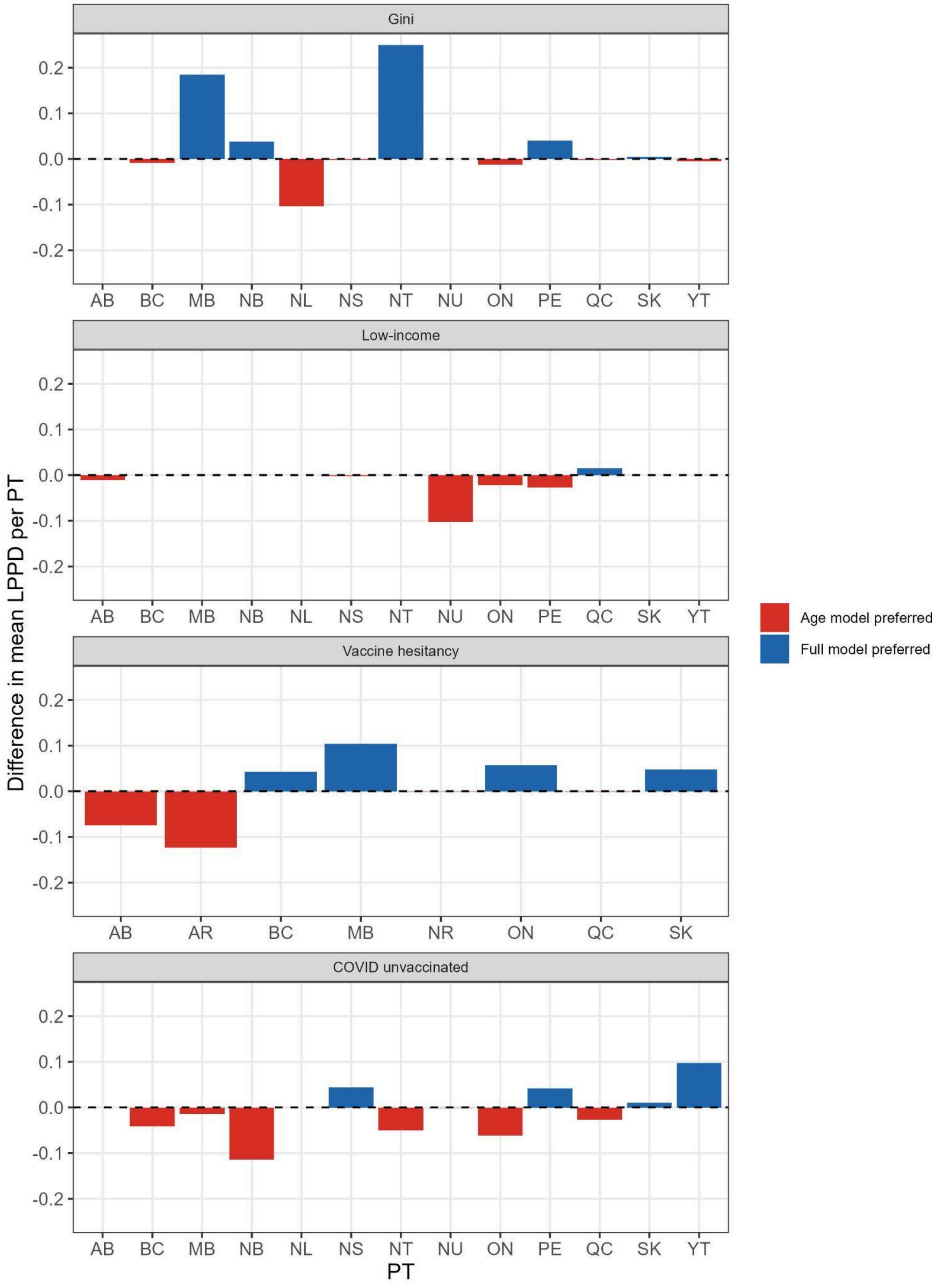

We saw that in our Gini and Vaccine Hesitancy modelling frameworks, the full (age+PT) model was preferred to the age-only proxy model. This matched the intuition gained from the best hyperparameters from model selection (Table 4 main text) wherein l2 was not maximal. In the Low Income and COVID-19 Unvaccinated modelling frameworks, relative differences in Supplementary Figure S2 were smaller with age-only model preferred overall, but note that the full model both frameworks already had maximal or near maximal l2 (constrained to 250 and 5 respectively by our plausible range). In other words, the Low Income modelling framework already favoured a predominantly age-centric model so was similar to the age-only proxy (cf. l2 = 250 and l2 = 500).

A natural further extension to this analysis would be to leave out groups of PTs (e.g. all territories or all Atlantic provinces); however, with only 13 PTs in total, leaving out groups of 3-4 simultaneously would leave limited training data and likely undesirable model fits. This would be a point for future work if/when richer data becomes available.

#### Simulation-based validation

Simulation-based validation provided an orthogonal way to evidence or inform validation of our model. The goal was to create a synthetic dataset with known fixed hyperparameters, run through the model, and see if our modelling selection process could recover the correct hyperparameter values. The procedure was as follows:

> Select values for the five model hyperparameters k, l1, l2, b, $\psi$, chosen randomly from the plausible search range defined above. From this we constructed a covariance matrix C and took one random draw from the multivariate normal distribution (i.e. our Gaussian Process prior model) with mean 0 and variance equal to C. We treated this draw as our synthetic "data". We then ran a proportion[2] of the data through our whole LOO-CV model selection pipeline and observed how well the hyperparameter values were recovered. We repeated this process 50 times.

Results are shown in Supplementary Table S2 with $\psi$ = 0. Our model selection was often very close to recovering the true parameter values. The 'normalized score' column ranks the predicted parameters' lppd, $lppd_{pred}$, against the true parameters' lppd, $lppd_{max}$, using the following formula: normalized score = ($lppd_{pred}$ - $lppd_{min}$) / ($lppd_{max}$ - $lppd_{min}$). We also report model performance using the full synthetic dataset each time as a comparison (denoted 'partial' and 'full' for the proportion of synthetic data used in model fitting).

[2] The partial synthetic dataset was restricted to the age-PT pairs present in the real data, ensuring that synthetic model selection operated on the same inferential problem as in the main model.

***<u>Supplementary Table S2:</u> Simulation-based validation: Our synthetic data used in simulation-based validation, together with summary results from our model selection process on each synthetic dataset.*** *Here $\psi$ = 0. The column 'Exact recovery?' shows in which synthetic datasets the model selection process was able to correctly recover all parameter values. The column 'Normalized lppd score' gives the relative performance in lppd between the predicted parameter values* ($lppd_{pred}$) *and true parameter values* ($lppd_{max}$)*, using the formula* normalized score = ($lppd_{pred}$ - $lppd_{min}$) / ($lppd_{max}$ - $lppd_{min}$). *Note that lppd for the worst-performing parameter combination in each case,* $lppd_{min}$, *was as large as* $-10^{12}$.

| Model framework | Relation of regions data | Vaccine coverage data |
|---|---|---|
| S | Gini indicator | Synthetic and normally-distributed with mean 0 and variance C. |

| Proportion of synthetic data used in model fitting | Number of iterations | Proportion of repetitions where the correct parameter value was retrieved | | | | | Mean normalized lppd score |
|---|---|---|---|---|---|---|---|
| | | k | l1 | l2 | b | all | |
| partial[2] | 50 | 0.98 | 0.38 | 0.46 | 0.64 | 0.12 | 0.92 |
| full | 50 | 1.00 | 0.48 | 0.44 | 0.70 | 0.22 | 0.91 |

| Data set (synthetic & partial) | True parameter values | | | | Predicted parameter values | | | | Exact recovery? | Normalized lppd score (2dp) |
|---|---|---|---|---|---|---|---|---|---|---|
| | k | l1 | l2 | b | k | l1 | l2 | b | | |
| 1 | exp | 1.50 | 0.50 | 1.00 | exp | 2.00 | 0.50 | 1.00 | false | 1.00 |
| 2 | sqexp | 1.00 | 2.50 | 1.50 | sqexp | 1.00 | 2.50 | 1.50 | true | 1.00 |
| 3 | exp | 1.75 | 1.50 | 1.50 | exp | 2.50 | 2.50 | 1.00 | false | 1.00 |
| 4 | sqexp | 1.00 | 0.50 | 1.50 | sqexp | 1.00 | 1.50 | 0.50 | false | 1.00 |
| 5 | sqexp | 2.50 | 0.20 | 1.00 | sqexp | 2.00 | 3.00 | 0.50 | false | 0.20 |
| 6 | sqexp | 1.75 | 4.00 | 1.50 | sqexp | 1.75 | 4.00 | 1.00 | false | 1.00 |
| 7 | exp | 2.00 | 3.00 | 0.50 | exp | 2.00 | 3.00 | 0.50 | true | 1.00 |
| 8 | sqexp | 1.75 | 0.20 | 0.50 | sqexp | 1.75 | 2.50 | 0.50 | false | 0.08 |
| 9 | exp | 2.50 | 3.50 | 0.50 | exp | 2.25 | 3.50 | 0.50 | false | 1.00 |
| 10 | exp | 1.50 | 1.50 | 1.50 | exp | 1.75 | 2.00 | 1.50 | false | 1.00 |
| 11 | exp | 2.50 | 1.00 | 1.50 | exp | 2.00 | 1.50 | 1.00 | false | 1.00 |
| 12 | sqexp | 1.25 | 2.50 | 1.00 | sqexp | 1.25 | 2.50 | 1.50 | false | 1.00 |
| 13 | sqexp | 1.00 | 10.00 | 1.00 | sqexp | 1.00 | 10.00 | 1.00 | true | 1.00 |
| 14 | sqexp | 2.25 | 10.00 | 1.50 | sqexp | 2.00 | 8.00 | 1.50 | false | 1.00 |
| 15 | exp | 1.50 | 2.50 | 1.50 | exp | 1.50 | 4.00 | 1.50 | false | 1.00 |
| 16 | sqexp | 2.00 | 1.50 | 0.50 | sqexp | 2.00 | 3.00 | 0.50 | false | 1.00 |
| 17 | sqexp | 2.00 | 25.00 | 1.50 | sqexp | 2.00 | 25.00 | 1.00 | false | 1.00 |
| 18 | exp | 1.50 | 25.00 | 1.50 | exp | 2.50 | 50.00 | 1.50 | false | 1.00 |
| 19 | sqexp | 1.00 | 3.00 | 1.00 | sqexp | 1.25 | 3.50 | 1.50 | false | 1.00 |
| 20 | sqexp | 1.75 | 0.20 | 1.50 | sqexp | 1.75 | 2.50 | 1.00 | false | 0.29 |
| 21 | exp | 1.00 | 0.20 | 1.50 | exp | 2.00 | 0.50 | 1.50 | false | 1.00 |
| 22 | sqexp | 1.75 | 3.00 | 0.50 | sqexp | 1.75 | 3.00 | 0.50 | true | 1.00 |
| 23 | exp | 1.50 | 2.50 | 0.50 | exp | 2.00 | 3.00 | 0.50 | false | 1.00 |
| 24 | exp | 2.50 | 10.00 | 1.00 | exp | 2.25 | 10.00 | 1.00 | false | 1.00 |

| | | | | | | | | | | |
|---|---|---|---|---|---|---|---|---|---|---|
| 25 | sqexp | 2.00 | 2.50 | 1.00 | sqexp | 2.00 | 3.00 | 1.00 | false | 1.00 |
| 26 | exp | 1.75 | 2.00 | 1.00 | exp | 1.00 | 2.50 | 0.50 | false | 1.00 |
| 27 | sqexp | 1.75 | 10.00 | 0.50 | sqexp | 2.00 | 10.00 | 1.00 | false | 1.00 |
| 28 | exp | 1.00 | 8.00 | 0.50 | exp | 1.75 | 10.00 | 0.50 | false | 1.00 |
| 29 | exp | 1.50 | 10.00 | 1.50 | exp | 1.75 | 25.00 | 1.00 | false | 1.00 |
| 30 | sqexp | 1.75 | 2.00 | 1.50 | sqexp | 1.75 | 2.50 | 1.00 | false | 1.00 |
| 31 | exp | 2.50 | 2.50 | 1.00 | exp | 1.25 | 1.50 | 1.00 | false | 1.00 |
| 32 | exp | 1.75 | 50.00 | 1.50 | exp | 1.25 | 50.00 | 1.50 | false | 1.00 |
| 33 | exp | 1.50 | 25.00 | 1.50 | exp | 1.25 | 25.00 | 1.50 | false | 1.00 |
| 34 | exp | 2.50 | 6.00 | 0.50 | sqexp | 1.00 | 10.00 | 0.50 | false | 1.00 |
| 35 | sqexp | 1.50 | 3.00 | 0.50 | sqexp | 1.50 | 3.00 | 0.50 | true | 1.00 |
| 36 | exp | 1.50 | 3.50 | 1.50 | exp | 1.00 | 2.50 | 1.00 | false | 1.00 |
| 37 | sqexp | 2.50 | 50.00 | 1.00 | sqexp | 2.00 | 50.00 | 1.00 | false | 1.00 |
| 38 | exp | 1.00 | 1.50 | 1.50 | exp | 1.00 | 2.00 | 1.50 | false | 1.00 |
| 39 | exp | 1.50 | 50.00 | 0.50 | exp | 2.00 | 50.00 | 0.50 | false | 1.00 |
| 40 | sqexp | 1.25 | 4.00 | 1.00 | sqexp | 1.25 | 4.00 | 1.00 | true | 1.00 |
| 41 | sqexp | 2.50 | 25.00 | 1.00 | sqexp | 2.25 | 25.00 | 1.00 | false | 1.00 |
| 42 | sqexp | 2.50 | 0.50 | 0.50 | sqexp | 2.00 | 3.00 | 0.50 | false | 0.41 |
| 43 | exp | 2.25 | 0.20 | 1.00 | exp | 1.50 | 0.20 | 1.00 | false | 1.00 |
| 44 | sqexp | 1.75 | 0.20 | 1.50 | sqexp | 1.75 | 2.50 | 0.50 | false | 0.02 |
| 45 | exp | 2.00 | 25.00 | 0.50 | exp | 2.25 | 25.00 | 0.50 | false | 1.00 |
| 46 | exp | 1.00 | 0.50 | 1.00 | exp | 1.25 | 0.50 | 1.50 | false | 1.00 |
| 47 | sqexp | 2.50 | 50.00 | 1.50 | sqexp | 2.00 | 50.00 | 1.00 | false | 1.00 |
| 48 | exp | 2.25 | 3.00 | 1.00 | exp | 2.25 | 3.50 | 1.00 | false | 1.00 |
| 49 | sqexp | 1.75 | 1.50 | 1.00 | sqexp | 2.00 | 2.50 | 1.50 | false | 1.00 |
| 50 | sqexp | 2.50 | 3.00 | 1.00 | sqexp | 2.25 | 3.00 | 1.00 | false | 1.00 |

#### Model testing - England comparison

To replicate the sparsity of Canada's data with the England data, we first restricted the refined Canadian data to the age range of the England data (6-17), then computed the 13 PT-level sample sizes. We then sampled 9/13 sample sizes (without replacement) and randomly assigned them to the 9 English regions. Finally, for each English region, we sampled the assigned number of vaccine coverage estimates to form the model training data; the remaining estimates acted as the test set. Since the same 9 Canadian PTs aren't selected each time, the sizes of the train and test sets vary. We repeated this process 40 times.

As an additional analysis, we repeated the above experiment, except without structuring the subsampling of the England data by region for training. Instead, we simply took 65 observations randomly from the England data per experimental replicate, matching the size of the Canada data for ages 6-17. This yields test sets that each have 43 observations in every replicate (108 total observations for England minus 65 training points).

For both experiments, we used the Gini index for total wealth from ONS 2016-2018 by region for the region relation indicator (Supplementary Table S3) [10] to complement the use of the Gini index for the Canadian data. Results for the first subsampling scheme are shown in the main text (Figure 2), while results for the second are in Supplementary Figure S3.

***Supplementary Table S3:*** ***Values used for the regions of England relation indicators.*** *See text for details. *NB: Very similar data values create an artificially strong relationship in our models. Modelling should be interpreted as a proof of concept only.*

| **Region of England** | **Region relation indicator** |
|---|---|
| | **Gini index (%)** |
| North East | 63.919 |
| East Midlands | 61.419 |
| Yorkshire and The Humber | 61.909 |
| West Midlands | 60.600 |
| South West | 58.981 |
| East of England | 58.942 |
| North West | 61.505 |
| South East | 57.815 |
| London | 69.601 |

### Supplementary results

***Supplementary Figure S3:*** ***Model accuracy against held-out test points when applied to England data subsampled to match the sparsity of the Canada data without regional structure.*** *Accuracy was quantified as the probability that the fitted model's predictions fall within 5% of each true (test) value. Hyperparameter optimization was performed anew for each subsampling. Panel A: aggregated distribution of accuracy scores (one score per test point) across all 40 subsamplings of the England data. The model achieved a higher mean accuracy than when the subsampling was additionally structured by region (88% vs 82%). Panel B: Model estimates (posterior draws; lines) from fitting the model to one subsampling (grey points), along with test data (points coloured by the corresponding accuracy score). Regions of England are shown as facets, ordered by their Gini index (ascending).*

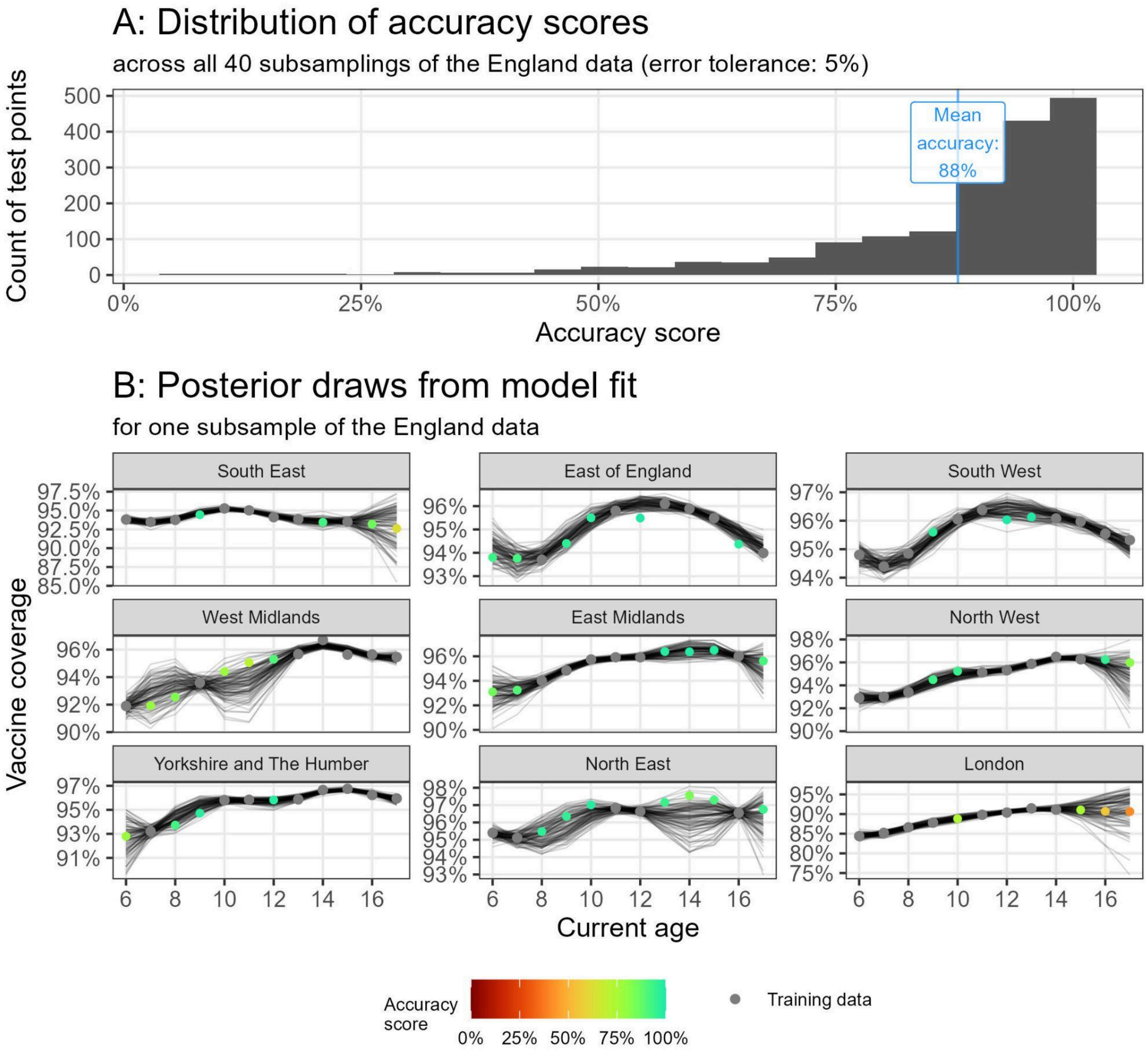

***Supplementary Figure S4:*** ***Visualizing the pairwise similarity in vaccine coverage based on the distance between PTs (Low Income indicator).*** *Figure elements are described in the Figure 3 caption (main text).*

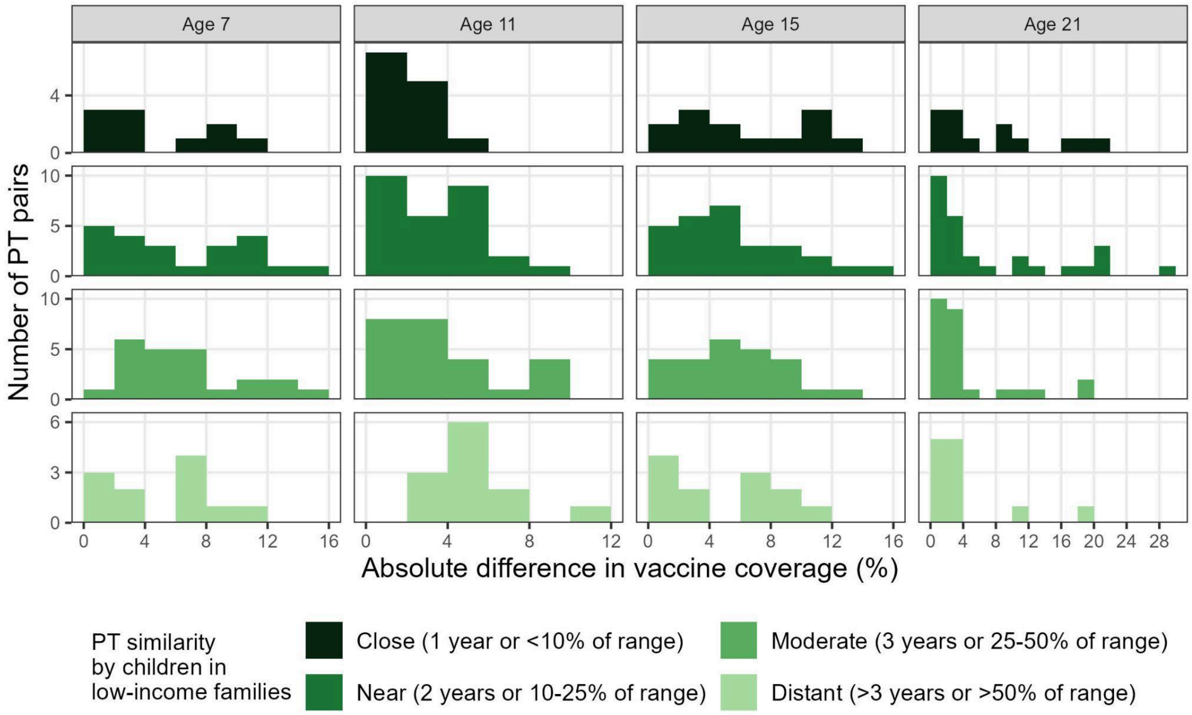


***Supplementary Figure S5:*** ***Visualizing the pairwise similarity in vaccine coverage based on the distance between PTs (Vaccine Hesitancy indicator).*** *Figure elements are described in the Figure 3 caption (main text).*

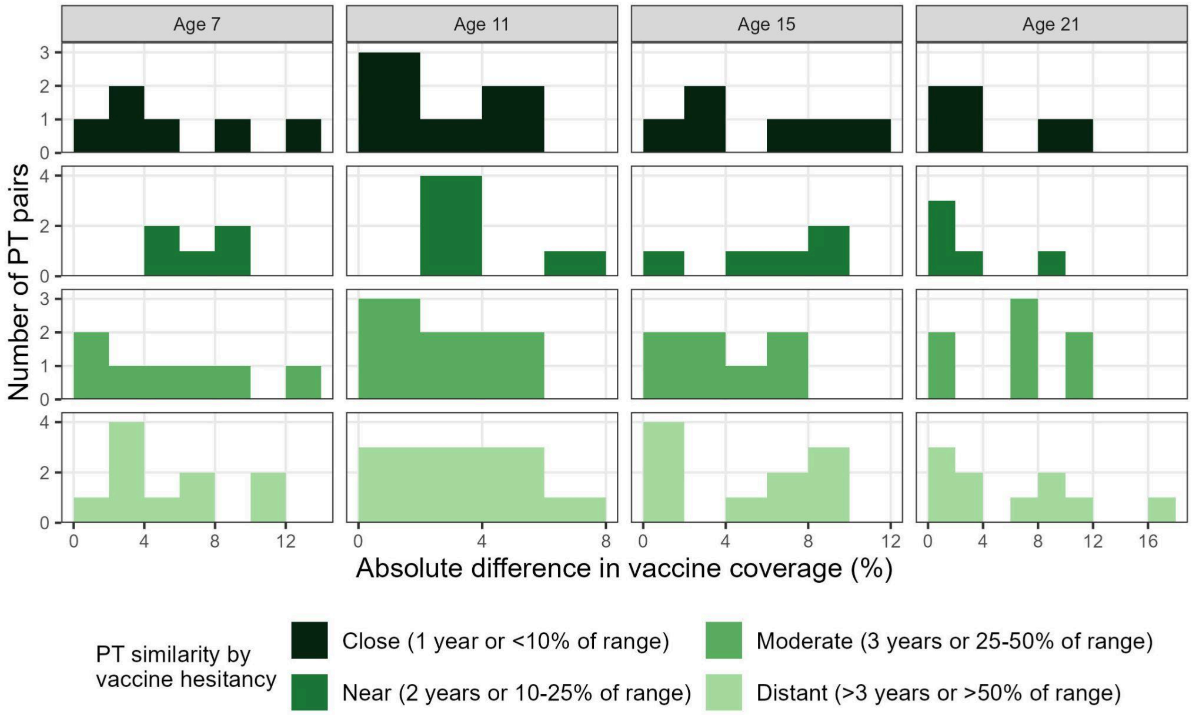

***<u>Supplementary Figure S6:</u>* Visualizing the pairwise similarity in vaccine coverage based on the distance between PTs (COVID-19 Unvaccinated indicator).** *Figure elements are described in the Figure 3 caption (main text).*

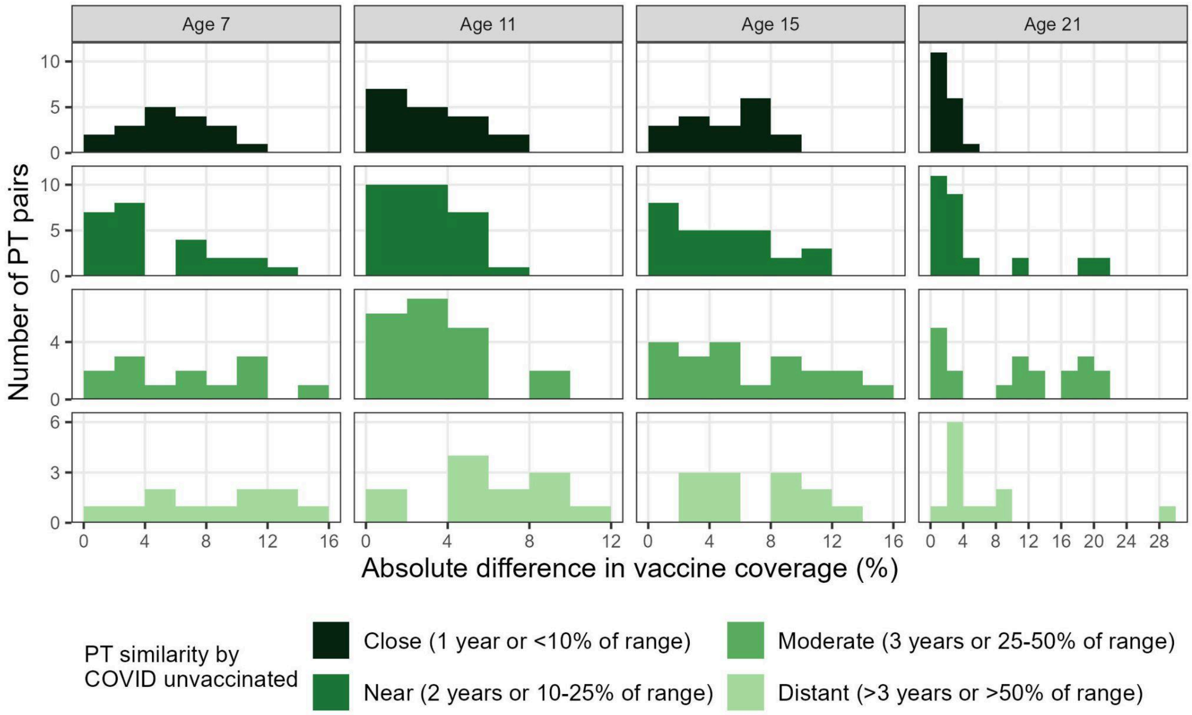

***Supplementary Figure S7:*** ***Model results for Canada showing estimates of vaccine coverage for at least one dose of a measles vaccine by age and province/territory (PT) for the Low Income model.*** *Black curves represent model estimates (posterior draws; 50 random draws). Grey points denote training data (1+ dose coverage). PTs are shown as facets, ordered by their Low Income indicator value (ascending). Model estimates are provided with a moderate-to-high degree of uncertainty, with estimates in smaller age gaps between observations typically associated with lower uncertainty. The comparison to 2+ dose data suggests plausible 1+ dose estimates, as we expect at least two dose coverage to be smaller than that of at least one dose.*

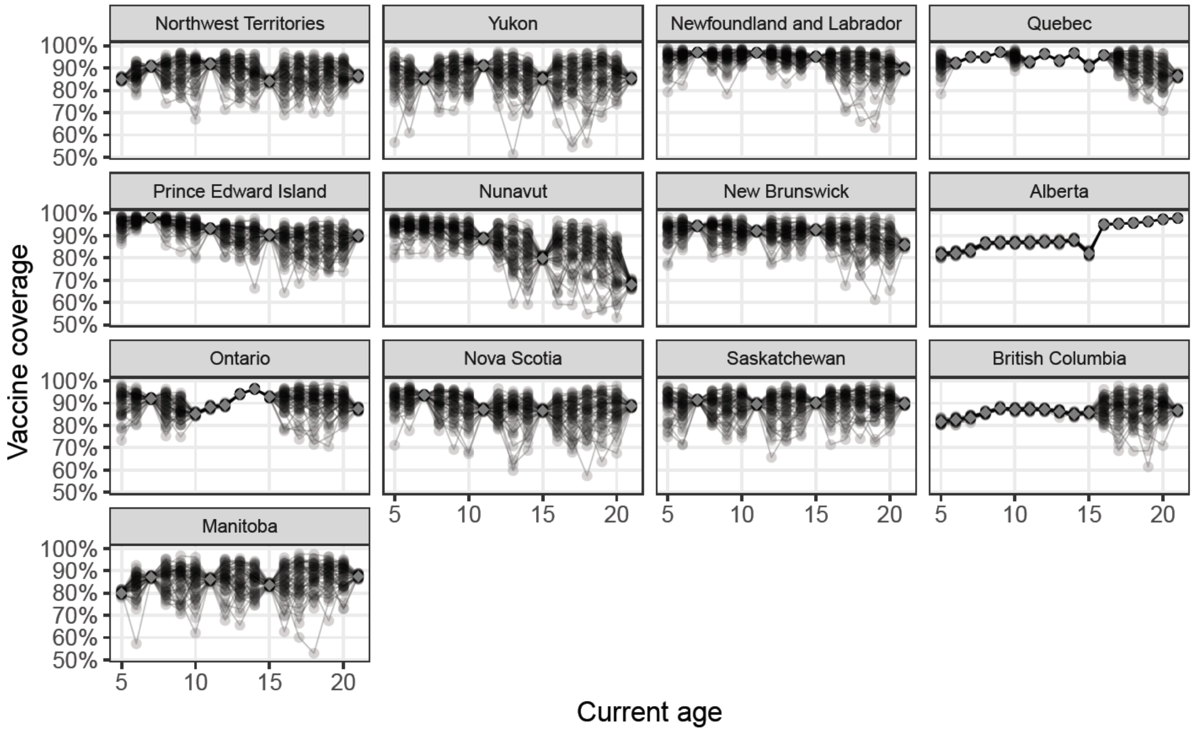

***Supplementary Figure S8:*** ***Model results for Canada showing estimates of vaccine coverage for at least one dose of a measles vaccine by age and province/territory (PT) for the Vaccine Hesitancy model.*** *Black curves represent model estimates (posterior draws; 50 random draws). Grey points denote training data (1+ dose coverage). PTs are shown as facets, ordered by their Vaccine Hesitancy indicator value (ascending). Model estimates are provided with a moderate-to-high degree of uncertainty, with estimates in smaller age gaps between observations typically associated with lower uncertainty. The comparison to 2+ dose data suggests plausible 1+ dose estimates, as we expect at least two dose coverage to be smaller than that of at least one dose.*

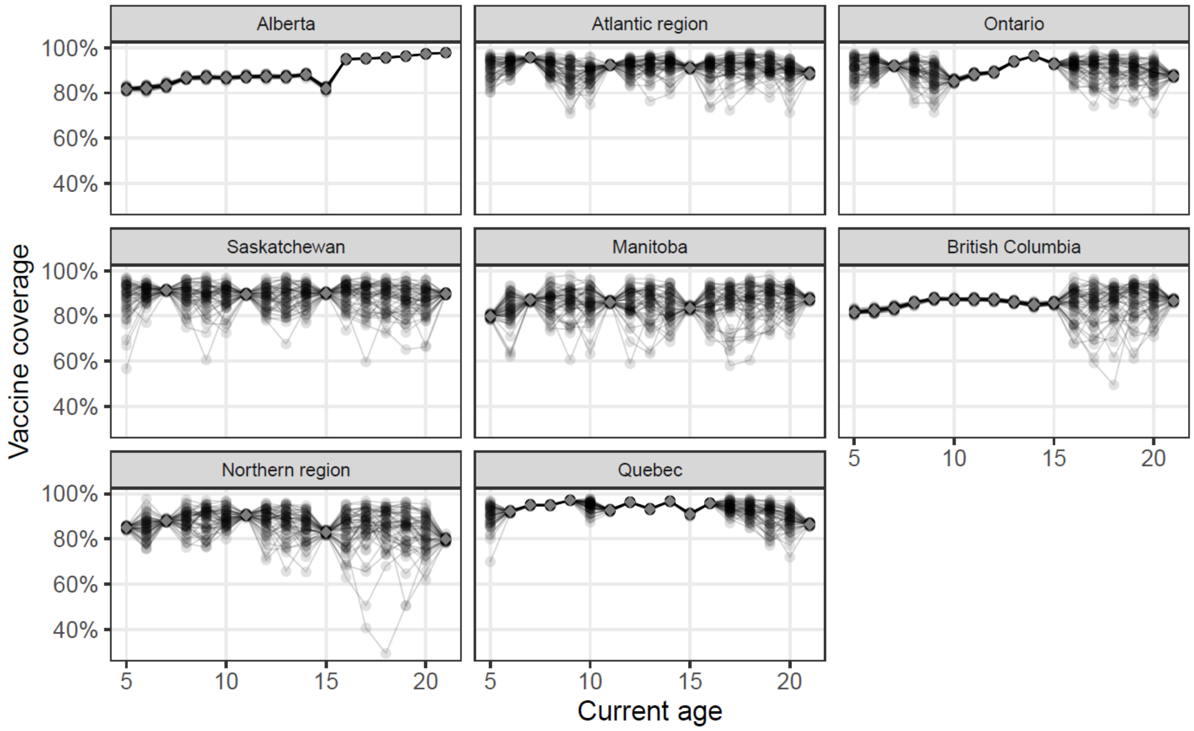

***Supplementary Figure S9:* Model results for Canada showing estimates of vaccine coverage for at least one dose of a measles vaccine by age and province/territory (PT) for the COVID-19 Unvaccinated model.** *Black curves represent model estimates (posterior draws; 50 random draws). Grey points denote training data (1+ dose coverage). PTs are shown as facets, ordered by their COVID-19 Unvaccinated indicator value (ascending). Model estimates are provided with a moderate-to-high degree of uncertainty, with estimates in smaller age gaps between observations typically associated with lower uncertainty. The comparison to 2+ dose data suggests plausible 1+ dose estimates, as we expect at least two dose coverage to be smaller than that of at least one dose.*

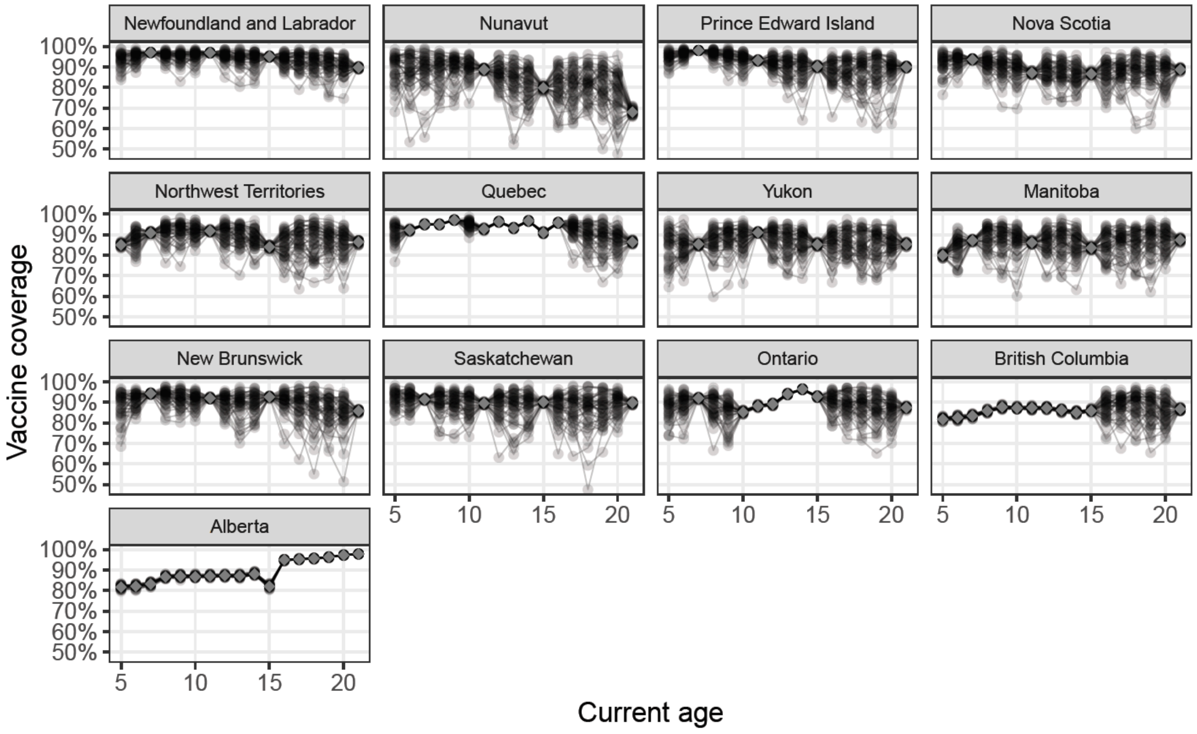

***<u>Supplementary Figure S10:</u>* Model results for Canada showing estimates of vaccine coverage for at least one dose of measles by age and province/territory (PT) - all modelling frameworks.** *Each row represents one modelling framework: the Gini model (top row), Low Income model (second row), Vaccine Hesitancy model (third row), and COVID-19 Unvaccinated model (bottom row). Black curves represent model estimates (posterior draws; 50 random draws) and red points denote training data (1+ dose coverage). PTs are shown as facets, ordered canonically. Model estimates are provided with a moderate-to-high degree of uncertainty, with estimates in smaller age gaps between observations typically associated with lower uncertainty. Each modelling framework uses the best-fit values from hyperparameter optimisation (Table 4) with measurement error* $\psi$ *= 0.05.*

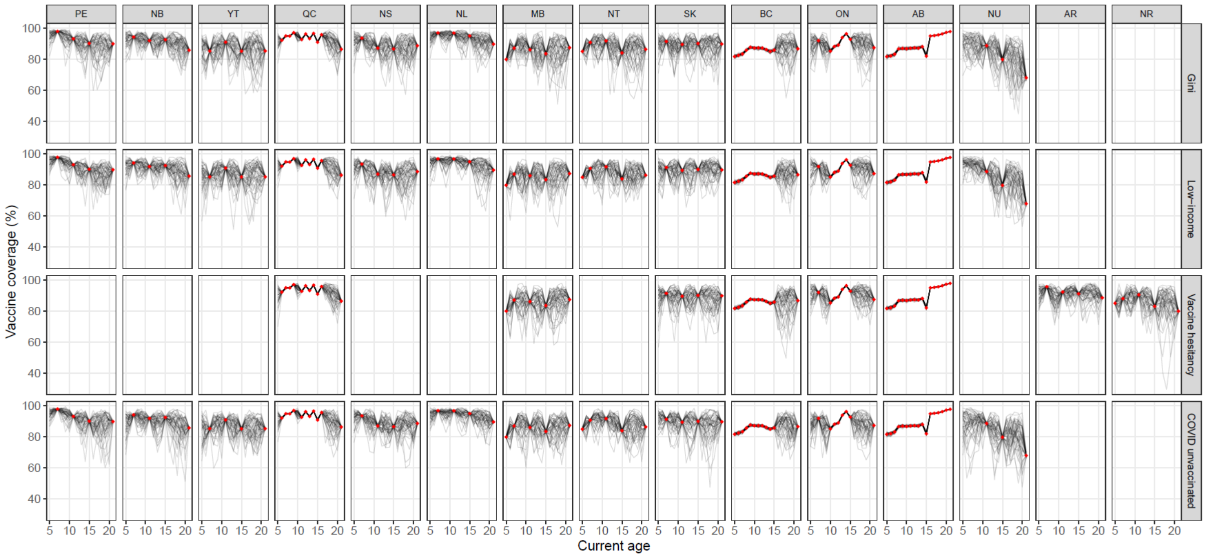
